# Primary Care Quality and Inappropriate Community Antibiotic Use: Evidence from Linked Provider and Household Data

**DOI:** 10.64898/2026.08.26.26361459

**Authors:** Yunwei Chen, Hongmei Yi, Sihang Rao, Ann Weber, Kristen Hassmiller-Lich, Sean Sylvia

**Author notes:** SS acknowledges support from the National Institutes of Health (K01AI159233, NIH/NIAID). HY acknowledges support from the Major Program of the National Social Science Fund of China (No. 23&ZD116), the “Health and Hope Fund” of the Business Development Center (BDC) of the Red Cross Society of China (RCSC) and UCB of Belgium. YC acknowledges support from the Doctoral Dissertation Fellowship from the University of North Carolina at Chapel Hill. We are grateful to Justin Trogdon, Marcos Vera-Hernandez, Grant Miller, and Anne Fitzpatrick for their insightful comments and suggestions. We also thank conference participants at the Rosenkranz Global Health Policy Research Symposium 2025 and the ASHEcon Annual Conference 2025, as well as seminar participants across various institutions for providing valuable input to this manuscript.

## Abstract

Inappropriate antibiotic use presents a major global health challenge, particularly in resource-limited settings where access to quality care remains limited while access to antibiotics is relatively unrestricted. This study estimates the relationship between frontline primary care quality and community antibiotic use, drawing on detailed data from approximately 100 rural villages in China that link objective measures of village doctors’ clinical practice quality, assessed through unannounced standardized patient visits, to household-level antibiotic use data measured within the same communities. To mitigate potential endogeneity concerns, we employed an instrumental variable (IV) approach integrated within a double/debiased machine learning (DML) framework, using ensemble machine learning to combine information across multiple individually weak but jointly informative provider-level instruments. Higher clinical practice quality was associated with lower probabilities of inappropriate antibiotic receipt during healthcare encounters for cold or diarrhea symptoms (OLS: 6.2 percentage points; DML-IV: 9.6 percentage points) and households storing antibiotics at home for future use (OLS: 4.1 percentage points; DML-IV: 10.9 percentage points). These findings highlight the potential role of improving frontline primary care quality in community antibiotic stewardship. Methodologically, this study illustrates how causal machine learning can complement conventional econometric methods by addressing high-dimensional estimation challenges in global health economics research.

## 1. Introduction

Clinically inappropriate use of antibiotics has become widespread across many low- and middle-income countries (LMICs) (Allwell-Brown et al., 2020; Fink et al., 2020; Klein et al., 2018; Rogawski et al., 2017). Although antibiotics are life-saving for bacterial infections, they are frequently used to treat respiratory illness and diarrhea, for which antibiotics are not recommended in most cases (Fink et al., 2020; Rogawski et al., 2017). This widespread misuse contributes to adverse effects on gut microbiota and immune systems, particularly in children, as well as the perpetuation of antimicrobial resistance at the population level (Jernberg et al., 2010; Johnson & Versalovic, 2012; Williams et al., 2018). Meanwhile, policies aimed at restricting inappropriate antibiotic use must avoid compromising access to effective treatment, making antibiotic stewardship a particularly challenging policy problem in low-resource settings (Laxminarayan et al., 2016; Okeke et al., 2024). In high-income settings, antibiotic access is typically regulated through prescription-only policies. However, such policies are considerably harder to implement in contexts where access to quality healthcare remains limited and frontline providers lack the ability to prescribe appropriately (Mendelson et al., 2016). In many LMICs, nonprescription antibiotics constitute a critical mechanism for accessing treatment among millions of people who struggle to obtain timely, high-quality healthcare (Morgan et al., 2011). A *Lancet* report highlighted that delayed or inadequate access to antibiotics causes more deaths than antibiotic resistance itself (Laxminarayan et al., 2016). Thus, although tightening prescription requirements may appear to be a straightforward response to inappropriate antibiotic use, such policies risk exacerbating undertreatment in areas where formal care is scarce or of low quality. Together, these competing considerations highlight a central policy challenge: how can inappropriate antibiotic use be reduced without undermining timely access to effective treatment in resource-limited settings?

Improving primary care quality is likely to play a key role in navigating this tradeoff. In many low-resource rural communities across LMICs, frontline primary care is delivered by village-level providers, who often have limited formal medical training but serve as the first, and sometimes only, point of contact for common illnesses (Arnold & Straus, 2005; Cox et al., 2017; Iwamoto et al., 2019). Their diagnostic and treatment practices influence not only whether patients receive antibiotics in clinical encounters, but also how households perceive illness severity and the value of antibiotics, potentially shaping antibiotic self-medication at home. Yet, evidence on how frontline primary care quality shapes community antibiotic use remains limited, despite community use accounting for 85-95% of global antibiotic consumption (Duffy et al., 2018). One central challenge is the scarcity of detailed community-based data linking primary care provider quality to antibiotic use within communities.

In this study, we examine the relationship between frontline primary care quality and community antibiotic use, focusing on a resource-limited setting where primary care quality remains limited while access to antibiotics is relatively unrestricted. We drew on detailed community-, provider-, and household-level data collected from approximately 100 rural villages in remote China, linking objective measures of village doctors’ clinical practice quality to household-level antibiotic use data collected from the same communities. Clinical practice quality was measured through unannounced clinical visits by standardized patients (SPs), who were recruited from local communities and presented a standardized disease case to village providers, enabling objective assessment of provider performance in real-world clinical encounters (Das & Hammer, 2014; Guo et al., 2020; Meng et al., 2022; Sylvia et al., 2015, 2017). We examined two complementary measures of community antibiotic use that capture antibiotic use through formal healthcare and household self-medication: antibiotic receipt during healthcare encounters for treating cold or diarrhea symptoms and household antibiotic storage as a proxy for self-medication practices.

To mitigate potential endogeneity concerns, we implemented an instrumental variable (IV) approach, leveraging extensive information collected from village doctors to construct candidate instruments for predicting their clinical practice quality. The rationale for instrument selection rests on the relatively stable placement of village doctors within their communities, which may limit provider sorting based on contemporaneous household health behaviors, and the use of upstream provider attributes that are conceptually motivated and plausibly predetermined relative to household antibiotic use. Nevertheless, causal interpretation of the IV estimates relies on instrument exogeneity and the exclusion restriction, requiring that the instruments affect household antibiotic use only through provider clinical practice quality. To assess the sensitivity of our findings to potential violations of the exclusion restriction, we additionally conducted plausibly exogenous IV bounding analyses under varying degrees of assumption relaxation (Conley et al., 2012).

To address a many-weak-instruments challenge, we integrated the IV approach within a double/debiased machine learning (DML) framework to accommodate high-dimensional data commonly encountered in community-based research (Chernozhukov et al., 2017, 2018). In this setting, no single candidate instrument strongly predicted the quality of provider clinical practice, limiting the ability of conventional IV models to satisfy the strong first-stage condition. We therefore incorporated multiple candidate instruments into a flexible DML-IV framework to approximate an optimal instrument by combining information across many individually weak but jointly informative instruments using an ensemble of machine learning algorithms. This approach allowed flexible modeling of high-dimensional instruments and controls, leveraging the rich provider-, household-, and community-level data collected in this setting, while preserving the interpretability of conventional IV estimation. The resulting instrument exhibited strong first-stage predictive performance, mitigating concerns about weak instruments.

We show that village doctors in this setting exhibited limited healthcare quality, yet remained the primary source of healthcare for families living in these communities. Antibiotic receipt for treating cold or diarrhea symptoms was common among facility users, and approximately half of households stored antibiotics at home, primarily for treating these conditions. Overall, our findings suggest that the clinical practice quality of village doctors is significantly associated with household antibiotic use. Specifically, a one-standard-deviation increase in the clinical practice quality index was associated with lower probabilities that patients received antibiotics during healthcare encounters for treating cold or diarrhea symptoms (OLS: 6.2 percentage points; DML-IV: 9.6 percentage points) and that households living in the same village stored antibiotics for future use (OLS: 4.1 percentage points; DML-IV: 10.9 percentage points). These findings were generally consistent across several robustness checks, while plausibly exogenous IV bounding analyses revealed varying degrees of sensitivity to potential violations of the exclusion restriction across outcomes.

To explore potential mechanisms underlying these relationships, we examined several pathways, including household healthcare-seeking behavior, antibiotic receipt conditional on visiting a village clinic, household perceptions of village clinic quality, and household antibiotic knowledge. Our exploratory analyses suggest that the relationship between primary care provider quality and household antibiotic use may operate primarily through differences in clinical decision-making and antibiotic prescribing during healthcare encounters (the intensive margin), rather than through changes in household care-seeking behavior (the extensive margin), with potentially important downstream implications for household antibiotic storage and self-medication. Specifically, we found little evidence of associations between provider quality and household healthcare-seeking behavior, village clinic utilization, perceived clinic quality, or antibiotic knowledge. In contrast, conditional on visiting a village clinic, higher provider quality was associated with a lower probability of patients receiving antibiotics for cold or diarrhea symptoms. Additionally, home-stocked antibiotics were commonly reported to originate from village clinics and pharmacies, suggesting that inappropriate antibiotic prescribing may contribute to the flow of unnecessary antibiotics into households, increasing the availability of leftover antibiotics available for future self-medication and potentially shaping subsequent medication-seeking behaviors when similar symptoms arise.

In a broad sense, this study contributes to the health economics literature on the relationship between provider quality and household health behaviors (Fe et al., 2017; Gauthier & Wane, 2011; Leonard, 2007, 2009). A long-standing literature on provider–patient information asymmetry emphasizes that patients often face substantial difficulty observing or evaluating provider quality, potentially limiting the extent to which differences in quality translate into household healthcare decisions (Rochaix, 1989). Another strand of the literature suggests that repeated interactions with providers may allow patients to learn about provider practices over time and adjust their health behaviors accordingly (Corno, 2014; Leonard, 2007; Leonard et al., 2009). Our findings highlight an important distinction between these channels. We find little evidence that objectively measured clinical quality substantially changes where households seek care, while higher clinical practice quality among frontline providers is associated with lower antibiotic receipt during clinical encounters, with potentially downstream implications for household antibiotic storage and subsequent self-medication or medication-seeking behaviors. In settings such as ours, where village doctors are deeply embedded in their communities and interact repeatedly with local households, improvements in clinical practice may shape household self-medication behavior through repeated treatment experiences even without substantial changes in provider choice. More broadly, our findings suggest that the relevance of frontline clinical quality may extend beyond individual healthcare encounters to downstream household health behavior.

This study also contributes to the growing literature on causal machine learning in applied economic research. Causal machine learning integrates flexible machine learning methods with causal inference frameworks to estimate parameters of interest in settings with complex and high-dimensional data (Athey & Imbens, 2019; Feuerriegel et al., 2024; Mullainathan & Spiess, 2017). While DML has been increasingly used to flexibly estimate nuisance functions and accommodate high-dimensional controls (Baiardi & Naghi, 2024; Shen & Zhang, 2024; Wichmann & Moreira Wichmann, 2023), our study illustrates a complementary application, using machine learning to leverage predictive information distributed across multiple candidate instruments within an IV framework when individual instruments provide limited first-stage predictive power. More broadly, this application demonstrates how causal machine learning can complement traditional econometric methods by leveraging rich, multidimensional data to address high-dimensional estimation challenges. This approach is particularly relevant to global health economics research, where increasingly rich data offer new analytical opportunities while also presenting complex estimation challenges.

This study has important policy implications for antibiotic stewardship in resource-constrained healthcare systems. Our findings suggest that strengthening frontline primary care quality may complement antibiotic stewardship strategies that primarily focus on restricting antibiotic use. Such efforts may be particularly relevant in resource-limited settings where community providers constitute the principal source of healthcare despite limited formal training and diagnostic capacity, and where restricting antibiotic access alone could unintentionally create barriers to appropriate treatment. Improving clinical practice quality, whether through conventional capacity-building and supervision or emerging approaches such as clinical decision-support systems and point-of-care diagnostic tools, may offer a promising strategy for strengthening community antibiotic stewardship in settings where access to quality healthcare remains limited while access to antibiotics is relatively unrestricted (Amin et al., 2023; Shapiro Ben David et al., 2025).

## 2. Institutional Background and Conceptual Framework

This study focuses on remote rural communities in China, a country characterized by high antibiotic consumption, relatively unrestricted over-the-counter access to antibiotics, and limited access to high-quality primary care in rural areas (Duan et al., 2021; Guo et al., 2020; Meng et al., 2022; Sylvia et al., 2015, 2017). China’s rural healthcare delivery system operates through a three-tier structure consisting of village clinics, township health centers, and county hospitals (Chen et al., 2022). Village clinics provide frontline primary care in villages, while township health centers and county hospitals function as higher-level facilities that supervise village clinics and manage referred patients. Township health centers and county hospitals may also provide primary care services directly, because patients are free to seek care at any level of the healthcare system in this setting. However, in many remote rural areas, such as this study setting in Yunnan Province in southwestern China, village clinics remain the primary source of primary care services due to geographical remoteness. As a result, the clinical practices of village providers may play a central role in shaping household antibiotic use in these communities.

Inappropriate community antibiotic use can be driven by both demand- and supply-side factors. On the demand side, studies from multiple countries have found substantial knowledge gaps among families regarding antibiotic use, high expectations for antibiotic effectiveness among patients, and widespread self-medication in settings where antibiotics are readily available without a prescription (Abdulah, 2012; Cantarero-Arévalo et al., 2017; Karras et al., 2003; Lin et al., 2020; Mangione-Smith et al., 1999; Szymczak et al., 2018; Yu et al., 2014). On the supply side, excessive and inappropriate prescribing is common among community providers in LMICs, driven possibly by limited clinical knowledge, diagnostic uncertainty, or inappropriate financial incentives (Currie et al., 2011; Di Martino et al., 2017; Garg et al., 2014; King et al., 2022; Li et al., 2012; Sun et al., 2015; Xue et al., 2018). Evidence from rural China suggests that deficits in diagnostic ability and uncertainty are major contributors to inappropriate antibiotic prescribing among village doctors (Kadirhaz et al., 2024; Xue et al., 2018).

Against this backdrop, the effect of primary care quality on community antibiotic use is theoretically ambiguous. The conceptual framework (Figure 1) illustrates how primary care quality may affect household antibiotic use through both the extensive and intensive margins of healthcare utilization, with additional downstream consequences for household self-medication. Along the extensive margin, higher-quality village doctors may influence whether and where households seek care, potentially shifting care towards village clinics or away from alternative sources such as higher-level healthcare facilities, pharmacies, or self-treatment (Fe et al., 2017; Gauthier & Wane, 2011; Leonard, 2007). Along the intensive margin, conditional on a healthcare encounter, higher clinical practice quality may improve diagnostic accuracy and treatment decisions, thereby reducing unnecessary antibiotic prescribing (Xue et al., 2018). These margins may operate in opposing directions. For example, better clinical quality may reduce antibiotic prescribing through improved diagnostic and treatment decisions, while changes in care-seeking may increase or decrease overall antibiotic use depending on differences in antibiotic-dispensing intensity across the healthcare sources between which patients shift.

**Figure 1.**
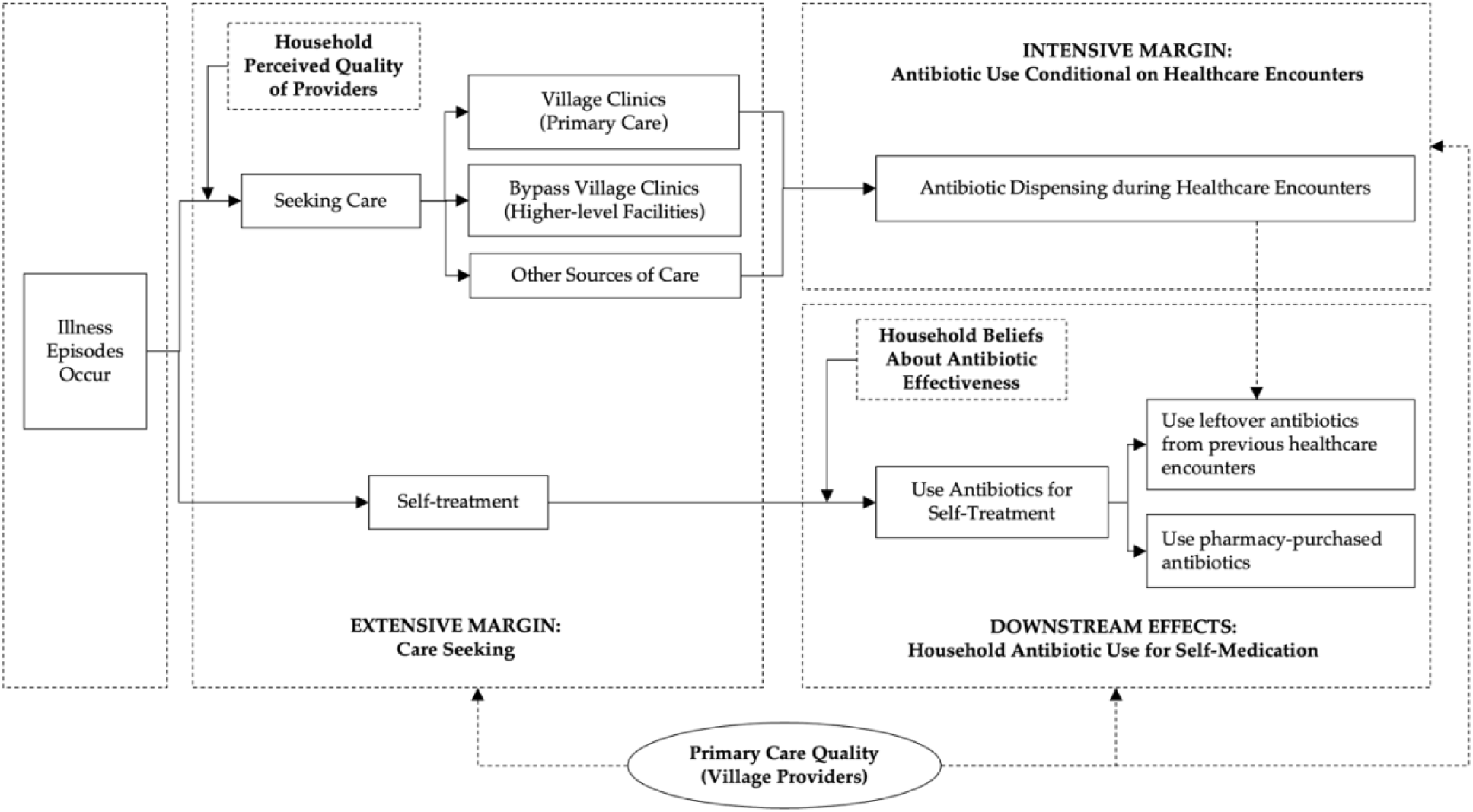
Conceptual Framework. *Notes:* This figure presents the conceptual framework that illustrates the various pathways through which primary care quality may affect household antibiotic use.

Importantly, patient-perceived quality may not coincide with objectively measured clinical quality. Providers’ incentives to accommodate patient demand for antibiotics may create a wedge between the objectively measured clinical practice quality and the quality perceived by patients, with the latter potentially shaping provider choice.

Provider quality may also have downstream effects beyond the immediate healthcare encounter. More appropriate prescribing may reduce the quantity of unnecessary antibiotics entering households and subsequently available for storage and future self-medication. Moreover, repeated interactions with providers may shape household beliefs, expectations, and trust regarding antibiotics and healthcare providers, potentially influencing future medication-seeking and healthcare-seeking behaviors (Ding et al., 2015; Mijović et al., 2022). Because these pathways may reinforce or offset one another, the effect of primary care quality on community antibiotic use remains ambiguous ex ante and depends on the relative magnitudes of these potentially offsetting channels. This ambiguity motivates our empirical examination of both antibiotic use during clinical encounters and antibiotic self-medication within households.

## 3. Data

### 3.1 Data Sources

The data were drawn from a research project conducted among rural clinicians in Yunnan Province in southwest China (Liu et al., 2022; Rao et al., 2021; Sylvia & Yi, 2017). A total of 330 villages were randomly selected from ten counties across three prefectures within Yunnan Province. Each village was represented by one village doctor, resulting in a study cohort of 330 village doctors.

In July 2017, comprehensive survey components were conducted across 330 villages. A clinic-level survey collected information on village and clinic characteristics, including geographical location, medicine inventory, and available medical equipment. A clinician-level survey captured provider characteristics such as demographics, qualifications, experience, recent training, and workload. In addition, all village doctors completed standardized clinical vignettes for three hypothetical cases (cold, diarrhea, and asthma) designed to assess their diagnostic and treatment knowledge. In these vignette assessments, trained enumerators portrayed standardized hypothetical patients presenting with hypothetical cases, and village doctors were asked to diagnose, prescribe, and manage each case as they would in routine clinical practice (Das & Hammer, 2005; Sylvia et al., 2015, 2017).

In December 2017, all 330 villages received an unannounced clinic visit from a SP presenting a standardized disease case to evaluate providers’ actual clinical practice. Unannounced SP visits are widely regarded as the gold standard for measuring quality of care (Das et al., 2008; Das & Hammer, 2014). SPs were recruited from local communities and trained to present standardized symptom profiles to village doctors to observe their real-world clinical behavior. While clinical vignettes primarily assess the upper bound of provider clinical knowledge, unannounced SP visits capture how providers actually diagnose and treat patients in routine practice.

In January 2018, a random subsample of approximately one-third of the villages (114) was selected for a household survey. Within each sampled village, 6-8 households were randomly chosen, yielding 692 households and 2503 individuals surveyed. The household-level survey collected basic demographic and socioeconomic information and detailed data on healthcare-seeking behavior and antibiotic use for each household member during episodes of common diseases, including cold or diarrhea symptoms in the previous year. In addition, trained enumerators systematically assessed each household’s medicine inventory and recorded the names, sources, and intended use of all antibiotics stored at home.

Among the 114 villages with household data, 103 successfully received an SP visit presenting a standardized asthma case, and the remaining villages failed to receive an SP assessment. We therefore restricted the analytical sample to these 103 villages with complete household and SP data and the 626 households and 2261 individuals associated with these villages.

### 3.2 Clinical Practice Quality of Village Doctors

We used data collected from unannounced SP visits to measure the clinical practice quality of village doctors. During these visits, SPs presented a standardized asthma case by mimicking symptoms typical of asthma patients. Because SP visits are logistically intensive and the repeated use of multiple cases increases the risk of SPs being recognized, it was feasible to implement only one standardized clinical case per village. The asthma case provides a clinically meaningful, guideline-based scenario that is broadly informative about provider clinical practice quality. First, asthma is an important primary care condition that requires careful history-taking and examination for accurate diagnosis and management, making it a strong test of providers’ real-world clinical practice. In addition, antibiotics are generally not indicated for uncomplicated asthma, so this case provides a clean setting in which inappropriate antibiotic prescribing can clearly be identified.

The unannounced SP visits followed a standard protocol. Each SP visited the village doctor without prior notice and initiated the consultation by stating, “Doctor, I feel short of breath,” then waited for the providers’ response. SPs were trained to provide consistent descriptions of symptoms such as wheezing, shortness of breath, and relevant triggers (e.g., exposure to allergens or physical exertion), but they only revealed these details if the provider asked appropriate questions.

Following each visit, SPs and trained enumerators recorded the questions the provider asked, the examinations performed, the diagnosis and treatment given, and all drugs prescribed. These elements were then compared with a checklist of recommended questions, examinations, and guidelines for asthma established by clinical experts. We used these data to construct three dimensions of village doctor clinical practice quality: (1) process quality, capturing whether providers obtained key clinical information through appropriate history taking and physical examination; (2) diagnostic quality, indicating whether providers correctly or partially diagnosed asthma; and (3) disease management quality, assessing whether providers appropriately managed the condition, including treatment and medication recommendations.

We then synthesized these indicators into an overall clinical practice quality index for each village doctor using the generalized least squares (GLS) weighting procedure, excluding items with zero responses (Anderson, 2008). This approach constructed a weighted average of standardized indicators, using weights derived from the inverse covariance matrix. The primary advantage of this procedure is that it accounts for correlations across individual indicators, down-weighting highly correlated indicators and giving more weight to indicators representing unique information. As a result, the index provides a more informative summary measure of each provider’s overall clinical practice quality, comparing to a summary or average score that counts each indicator equally. The resulting quality index was approximately normally distributed (Figure S1) and was analyzed as a continuous measure of clinical practice quality.

### 3.3 Household Antibiotic Use

To capture household antibiotic use comprehensively, we examined two complementary measures. First, we measured antibiotic receipt among patients seeking treatment for cold or diarrhea symptoms at a healthcare facility, capturing antibiotic use during clinical encounters. Because this outcome is observed only among facility users, we therefore also examined household antibiotic storage as a proxy for antibiotic use through home self-medication.

The first outcome measured antibiotic receipt during healthcare encounters among individuals who sought treatment for cold or diarrhea symptoms. During the household survey, respondents reported whether each household member experienced an episode of cold or diarrhea symptoms during the previous year, whether formal treatment was sought, and the type of healthcare facility visited. Among individuals who sought formal care, respondents were asked whether antibiotics were prescribed during that visit. We coded this as a binary outcome indicating whether the individual received antibiotics for the reported illness episode during the clinical encounter (1 = yes, 0 = no).

In contrast, the second outcome, household antibiotic storage, is an objective measure obtained during the household survey. Trained enumerators visually assessed each household’s medicine inventory and recorded the names, sources, and intended uses of all antibiotics stored in the home. We defined a binary outcome indicating whether the household stored any antibiotics (1 = yes, 0 = no), as an objective proxy for antibiotic availability for potential self-medication practices within the household.

Together, these two complementary outcomes capture the two principal pathways of community antibiotic use and are measured at different levels. Antibiotic prescribing for treating cold or diarrhea symptoms during healthcare encounters is measured at the individual illness-episode level among individuals who seek treatment, whereas household antibiotic storage is measured at the household level as an objective proxy for home self-medication practices.

## 4. Empirical Strategy

### 4.1 Baseline OLS Specification

In the baseline OLS specification, we began by specifying the relationship between village doctor clinical practice quality and household antibiotic use as follows:

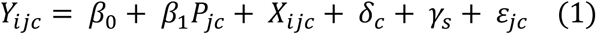

where *Y_ijc_* denotes the antibiotic use outcome for observation *i* in village *j* of county *c.* Depending on the outcome, the unit of observation *i* is either an individual illness episode, for antibiotic receipt during healthcare encounters, or a household, for household antibiotic storage. *P_jc_* denotes the clinical practice quality index for the village doctor serving village *j* of county *c*. *X_ijc_* includes covariates at the village, household, individual, and illness-episode levels. Village-level controls included population size, remoteness, access to alternative clinics and pharmacies, and provider nativeness and practice in the current village. Household-level controls included household size, presence of young children, and household wealth index. For the individual illness-episode outcome, we further adjusted for individual characteristics (age, gender, and health status index) and included illness-type fixed effects (cold vs. diarrhea) to account for systematic differences in healthcare-seeking and treatment across illness types. We also included county fixed effects (*δ_c_*) to account for unobserved county-level heterogeneity at the higher administrative level and SP fixed effects (*γ_s_*) to account for potential differences across standardized patients. Standard errors are clustered at the township level.

### 4.2 IV Estimation

Because provider quality is not randomly assigned across villages, the OLS estimates may be biased by unobserved differences in healthcare demand, provider selection, and other factors correlated with both provider quality and household antibiotic use. To mitigate potential endogeneity concerns, we complemented the OLS analyses with an IV framework (Figure S2).

#### 4.2.1 Candidate Instruments

We constructed a set of candidate instruments from detailed provider-level data collected during the village clinician survey. This instrument set comprises 18 provider characteristics spanning four domains (Table S1): (1) demographics (age, gender, ethnicity); (2) education and credentials (years of medical practice, education level, formal medical diploma, and medical practice license); (3) recent training exposure (training at township, county, city, or online levels during the previous year); and (4) vignette-based clinical competence, which proxies latent clinical ability. Clinical vignettes were administered during the village clinician survey using three hypothetical cases (asthma, cold, and diarrhea; Tables S2-S4). Responses to the clinical vignettes were used to construct provider competence indices following the same GLS weighting procedure as for the clinical practice quality index, including an overall competence index, three disease-specific indices (asthma, cold, and diarrhea), and three domain-specific indices (process, diagnostic, and treatment competence).

#### 4.2.2 Instrument Validity

The exclusion restriction is a crucial assumption for these proposed candidate instruments to be valid. That is, the candidate instruments are uncorrelated with unobserved determinants of household antibiotic use and affect outcomes only through provider clinical practice quality. Although these assumptions cannot be directly tested, we consider several features of the study setting and instrument selection that may support their plausibility.

First, the selection of candidate instruments rests on upstream provider-level attributes that are plausibly predetermined relative to household health behavior and may influence household antibiotic use primarily through provider clinical practice quality. They include demographic characteristics (e.g., age), historically established credentials (e.g., medical practice license), independently assessed clinical competence (e.g., vignette-based competence), and training exposure that may be influenced by institutional policies. We excluded provider attributes more likely to be directly related to contemporaneous household healthcare demand, such as patient volume or working hours, although these measures were available in our data.

Second, village doctors exhibited long-term, stable placement within their communities (Figure S3). Village doctors had practiced in their current villages for an average of 15.7 years, with over 75% practicing in the same village for more than 10 years. This relatively limited mobility may reduce concerns about provider sorting based on contemporaneous household health behaviors.

Third, we adjusted for observed village and provider characteristics to address potential alternative pathways linking provider attributes to household antibiotic use, leveraging the rich contextual information collected in this study. First, to account for the possibility that more qualified providers were differentially allocated to villages with better infrastructure and healthcare access during early placement, we controlled for village-level characteristics, including population size, remoteness, distance to township and county seats, and access to alternative clinics and pharmacies. Second, to account for potential non-clinical interactions between providers and local households, we adjusted for provider community embeddedness using indicators of village nativeness and long-term practice in the current village.

Fourth, we included county fixed effects to control for systematic differences across counties, such as county-level regulation, healthcare resources, and institutional environments. This specification leverages within-county variation in provider clinical practice quality and household antibiotic use, reducing concerns about confounding from shared county-level characteristics. Thus, after controlling for village resources, provider community embeddedness, and county fixed effects, some variation in provider attributes may be plausibly considered conditionally random.

While these features may support the plausibility that the proposed provider attributes affect household antibiotic use primarily through their realized clinical practices, potential violations of the exclusion restrictions cannot be ruled out due to alternative pathways we could not account for in this setting. To assess the sensitivity of the IV estimates to potential violations of the exclusion restriction, we additionally employed the plausibly exogenous IV bounding approach proposed by Conley et al. (2012), which allows for bounded violations of the exclusion restriction and examines how the IV estimates and their confidence intervals vary under different degrees of assumption relaxation.

#### 4.2.3 Limitations of Conventional IV

A key challenge in this setting is that no individual candidate instrument strongly predicts their clinical practice quality, making it difficult to satisfy the strong first-stage condition required for IV estimation. Including all candidate instruments in a conventional first-stage regression is also undesirable because a large set of individually weak instruments can lead to imprecise estimates and weak-instrument bias.

Recent work has proposed machine learning approaches to approximate an “optimal” instrument from high-dimensional data, most commonly using lasso-based variable selection (Belloni et al., 2012; Chernozhukov et al., 2015a; Danquah et al., 2021). These methods perform well when the first stage is approximately sparse, that is, when a relatively small subset of instruments contains most of the predictive information (Belloni et al., 2014b, 2014a; Chernozhukov et al., 2015b). In our setting, however, predictive information is distributed across many provider characteristics. As a result, lasso selected too few instruments to recover a sufficiently strong first stage (Belloni et al., 2012). These limitations motivate a more flexible estimation strategy that can more efficiently aggregate predictive information across multiple candidate instruments while preserving the causal interpretation of the IV framework.

### 4.3 DML-IV Estimation

To address this many-weak-instruments challenge while leveraging the extensive provider information available in our data, we integrated the double/debiased machine learning (i.e., DML) approach within the IV framework described above (Chernozhukov et al., 2017, 2018). DML can flexibly combine information from many candidate instruments using a broad class of supervised machine learning methods, thereby strengthening first-stage prediction and aggregating information more effectively from many weak but jointly informative provider information (Ahrens et al., 2024, 2026; Chernozhukov et al., 2017, 2018).

Specifically, we implemented a DML-IV model that allows flexible modeling of high-dimensional instruments and controls. We used the 18 provider characteristics described above as the baseline instrument set for the DML procedure and included first-order interactions and second-order polynomial terms to approximates an optimal instrument using an ensemble of supervised machine learning algorithms, including OLS, cross-validated ridge, random forest, gradient boosting, and neural networks. We also implemented a recently developed stacking approach that combines multiple machine learners into a single meta-learner to improve first-stage prediction, reducing the reliance on any single machine learning algorithm (Ahrens et al., 2023, 2025). To account for within-township dependence, estimation used cluster-dependent cross-fitting, with standard errors clustered at the township level. Additional methodology details are provided in Appendix Text C.

## 5. Results

### 5.1 Descriptive Statistics

#### 5.1.1 Village and Healthcare Setting

Table 1 presents the characteristics of 103 villages. These study villages were sparsely populated and geographically remote. Half of the villages (50.5%) had fewer than 3,000 residents within a five-kilometer radius, with roughly 75% of villages located more than five kilometers from the township seat and nearly 80% situated more than 20 kilometers from the county seat.

**Table 1.** Summary statistics of villages (N=103).

|  | **n(%)** |
|---|---|
| **Villages** |  |
| Population within 5 km |  |
| <=3000 population | 52 (50.5%) |
| 3000-10000 population | 34 (33.0%) |
| >10000 population | 17 (16.5%) |
| Distance from village to township seat |  |
| <=5 km | 27 (26.2%) |
| 5-10 km | 22 (21.4%) |
| >10 km | 54 (52.4%) |
| Distance from village to county seat |  |
| <=20 km | 21 (20.4%) |
| 20-50 km | 42 (40.8%) |
| >50 km | 40 (38.8%) |
| Number of clinics within 5 km |  |
| Only one | 26 (25.2%) |
| 2-4 clinics | 44 (42.7%) |
| >=5 clinics | 33 (32.0%) |
| Number of pharmacies within 5 km |  |
| None | 46 (44.7%) |
| >=1 pharmacy | 57 (55.3%) |
| **Village Clinics** |  |
| Monthly patient volume |  |
| <=100 patients | 39 (37.9%) |
| 100-500 patients | 36 (35.0%) |
| >500 patients | 28 (27.2%) |
| Number of western drugs available last year |  |
| <=50 drugs | 42 (40.8%) |
| 50-100 drugs | 31 (30.1%) |
| >100 drugs | 30 (29.1%) |
| Number of Chinese patent drugs available last year |  |
| <=50 drugs | 80 (77.7%) |
| >50 drugs | 23 (22.3%) |
| Number of herbal drugs available last year |  |
| None | 73 (70.9%) |
| >=1 drug | 30 (29.1%) |

*Note.* Data are presented in n (%).

Village clinics constitute the primary source of frontline care in these remote communities. One-quarter of villages (25.2%) had access to only one clinic within five kilometers, and nearly half (44.7%) had no pharmacy within the same distance. More than 60% of village clinics reported over 100 patient visits per month, and nearly 30% reported more than 500 monthly visits. Village clinics primarily stocked Western medicines. Most village clinics (59.2%) stocked more than 50 Western medicines, whereas only 22.3% stocked more than 50 Chinese patent medicines. Herbal medicines were rarely stocked, indicating a predominant reliance on Western pharmaceutical treatments.

#### 5.1.2 Household Characteristics and Healthcare Access

Table 2 describes the characteristics of the 626 households surveyed across the 103 villages. The sample reflected the socioeconomic conditions typical of remote rural communities. More than 30% were registered as poverty-stricken households. Ownership of household assets was modest: while televisions (92.8%) and washing machines (77.2%) were common, fewer households owned refrigerators (52.9%), water heaters (35.3%), kitchen hoods (10.2%), modern cooking fuels (8.9%), or flush toilets (4.5%). Only 22.8% of household heads had completed at least a middle school education.

**Table 2.** Summary statistics of households (N=626).

|  | **Mean (SD) / n (%)** |
|---|---|
| Household size | 3.6 (1.7) |
| Household with children under ten (0/1) | 230 (36.7%) |
| Household head completed middle school (0/1) | 143 (22.8%) |
| Poverty-stricken household registered with the government (0/1) | 204 (32.6%) |
| Household assets |  |
| Owned a TV (0/1) | 581 (92.8%) |
| Owned a washing machine (0/1) | 483 (77.2%) |
| Owned a fridge (0/1) | 331 (52.9%) |
| Owned a water heater (0/1) | 221 (35.3%) |
| Owned a kitchen hood (0/1) | 64 (10.2%) |
| Owned a modern fuel (gas) (0/1) | 56 (8.9%) |
| Owned a flush toilet (0/1) | 28 (4.5%) |
| Distance |  |
| The village clinic is within 1 km (0/1) | 430 (68.7%) |
| The frequently-visited township health center is within 5 km (0/1) | 175 (28.0%) |
| The frequently-visited county hospital is within 20 km (0/1) | 123 (19.8%) |
| Household healthcare seeking preference |  |
| Home Self-medication | 168 (26.9%) |
| Pharmacy | 37 (5.9%) |
| Private clinics | 42 (6.7%) |
| Village clinics | 271 (43.4%) |
| Township health centers | 38 (6.1%) |
| County hospitals | 44 (7.1%) |
| City hospitals | 6 (1.0%) |
| Others | 18 (2.9%) |

*Note:* Data are presented in n (%) for binary variables and mean (SD) for continuous variables.

Access to primary healthcare by households was centered on village clinics. Nearly 70% of households reported having access to a village clinic within one kilometer. In contrast, township health centers (28% within five kilometers) and county hospitals (19.8% within 20 kilometers) were less accessible. When asked about the preferred healthcare provider when ill, village clinics were the most commonly chosen option (43.4%), followed by home self-medication (26.9%). Other choices, such as visiting a pharmacy or seeking care at higher-level facilities like township health centers or county hospitals, were much less common, each reported by fewer than 10% of households.

Households also viewed village clinics as the most cost-effective option for treating common illnesses. In the household survey, respondents were asked how many days and how much money they believed recovery would require if they sought care at each facility type for treating a cold or diarrhea condition. These responses were used to construct indices of perceived effectiveness and cost for each facility type. Figure S4 shows that households considered village clinics to be as effective as township health centers and more effective than county hospitals. At the same time, households perceived village clinics to be less costly than either township health centers or county hospitals (Figure S5). Together, they highlight the central role of village clinics in providing frontline primary care in these remote rural communities.

#### 5.1.3 Characteristics of Village Doctors

Table 3 describes the characteristics of the 103 village doctors serving these communities. Most village doctors were male (60.2%), with an average age of 45.2 years. They had practiced medicine for an average of 20.8 years, 15.7 of which were spent in their current village. Approximately 10% were ethnic minorities, and 78.6% were native to the villages where they practiced.

**Table 3.**
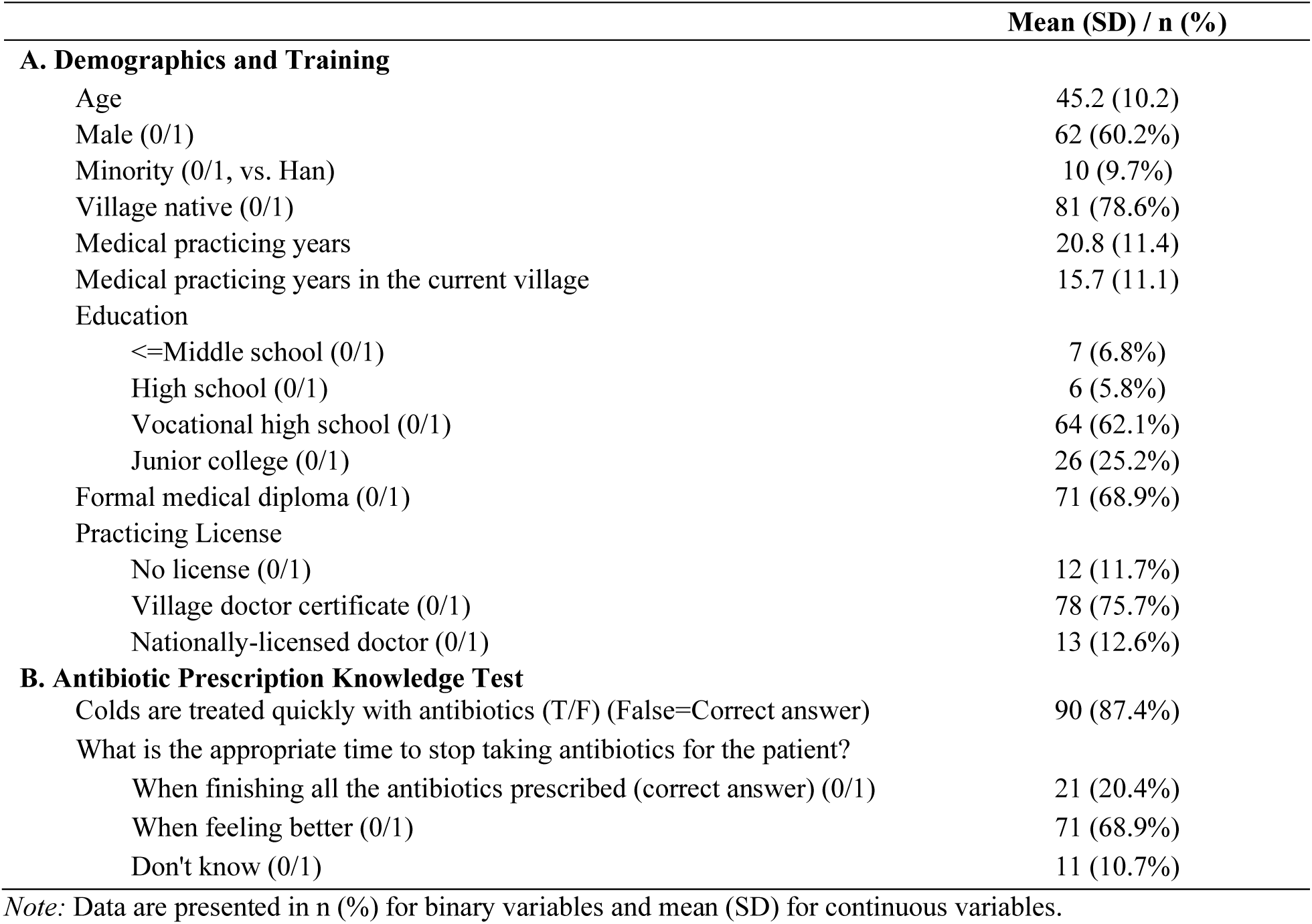
Summary statistics of village doctors (N=103).

Formal medical training and professional qualifications were limited. About two-thirds (67.9%) had completed only high school or vocational high school, 6.8% had no more than a middle school education, and 25.2% had completed junior college. None held a college degree or higher. While around 70% reported receiving some form of formal medical education, most (75.7%) practiced under a village doctor certificate rather than a national medical license. Village doctor certificates, typically issued to former barefoot doctors, village health workers, or traditional practitioners, authorize practice only in village clinics under the supervision of local health authorities. Only 12.6% held a national medical license, while 11.7% reported having no formal medical license.

Knowledge of appropriate antibiotic use among village doctors was limited. While most village doctors (87.4%) correctly recognized that the common cold could not be treated with antibiotics quickly, only 20.4% correctly identified that antibiotics should be discontinued only after completing the prescribed course. In contrast, 68.9% incorrectly believed that patients could stop taking antibiotics once symptoms improved, and 10% were uncertain. Together, these statistics indicate that these village doctors possessed strong community ties, but had limited formal education and training and showed substantial knowledge gaps regarding appropriate antibiotic use.

#### 5.1.4 Clinical Practice Quality of Village Doctors

Table 4 summarizes the clinical practice quality of village doctors based on unannounced SP visits presenting a standardized asthma case. Overall, village doctors exhibited substantial deficiencies across all three dimensions of clinical practice quality: clinical process, diagnostic accuracy, and treatment management.

**Table 4.** Summary statistics of clinical practice quality among village doctors (N=103).

|  | **n(%)** |
|---|---|
| **A. Recommended Questions for Diagnosing Asthma** |  |
| 1. Symptoms onset last time | 27 (26.2%) |
| 2. The progression of the disease | 4 (3.9%) |
| 3. Approaches to alleviating symptoms | 2 (1.9%) |
| 4. Triggers of symptoms | 18 (17.5%) |
| 5. Intensity and duration of symptoms | 11 (10.7%) |
| 6. Difficulty breathing | 7 (6.8%) |
| 7. Onset of first symptoms | 40 (38.8%) |
| 8. Sounds of breathing (wheezing) | 1 (1.0%) |
| 9. Cold/fever | 46 (44.7%) |
| 10. Coughing/expectoration | 61 (59.2%) |
| 11. Medical history of the family | 1 (1.0%) |
| 12. Other diseases | 35 (34.0%) |
| 13. History of childhood illnesses | 1 (1.0%) |
| **B. Recommended Exams for Diagnosing Asthma** |  |
| 1. Chest/back auscultation | 19 (18.4%) |
| 2. Pulmonary ventilation test | 0 (0.0%) |
| 3. Bronchodilator test (airway reversible test) | 0 (0.0%) |
| 4. Physical examination | 23 (22.3%) |
| 5. Chest X-ray test | 0 (0.0%) |
| 6. Routine blood tests | 0 (0.0%) |
| 7. Unvoiced double lung percussion | 0 (0.0%) |
| **C. Diagnosis** |  |
| Gave correct diagnosis | 5 (4.9%) |
| Gave correct or partially correct diagnosis | 11 (10.7%) |
| **D. Treatment** |  |
| Recommended avoiding allergens (dust and particulates) | 3 (2.9%) |
| Correct medication: inhaled/oral corticosteroids and beta-receptor agonists | 4 (3.9%) |
| No antibiotics prescribed | 82 (79.6%) |

*Note:* Data are presented as n (%). Village doctor clinical practice quality was measured using unannounced standardized patient visits presenting a standardized asthma case. Correct diagnoses include asthma or bronchial asthma**.** Partially correct diagnoses include dyspnea-related conditions without a specific asthma diagnosis.

Although some commonly recommended questions were frequently asked, such as coughing (59.2%), cold or fever symptoms (44.7%), or onset of first symptoms (38.8%), the majority of village doctors missed several critical questions for asthma diagnosis, such as disease progression (3.9%), symptom alleviation approaches (1.9%), sounds of breathing (1.0%), and childhood illnesses (1.0%). Diagnostic accuracy was notably low. Only five village doctors (4.9%) correctly diagnosed asthma, while an additional six recognized a breathing-related condition (e.g., dyspnea) without identifying asthma specifically. Treatment management quality was also limited. Only four village doctors (3.9%) prescribed appropriate asthma medication, and only three (2.9%) advised avoiding common environmental triggers, such as dust or airborne particulates. Although most village doctors (82, 79.6%) appropriately refrained from prescribing antibiotics for the asthma case, a nontrivial proportion still prescribed antibiotics despite failing to establish the correct diagnosis, suggesting potentially inappropriate antibiotic use.

Taken together, these findings reveal substantial deficiencies in the quality of frontline primary care provided by village doctors, a common challenge in rural and remote communities globally. At the same time, the quality index derived from these clinical practice quality indicators exhibited substantial variation across village doctors (Figure S2), indicating meaningful heterogeneity in provider quality across these study communities and providing the empirical basis for examining whether provider quality contributes to inappropriate household antibiotic use.

#### 5.1.5 Household Antibiotic Use

Table 5 summarizes the two primary measures of household antibiotic use in these rural communities, including antibiotic receipt during healthcare encounters and household antibiotic storage, serving as a proxy for antibiotic use through home self-medication.

**Table 5.**
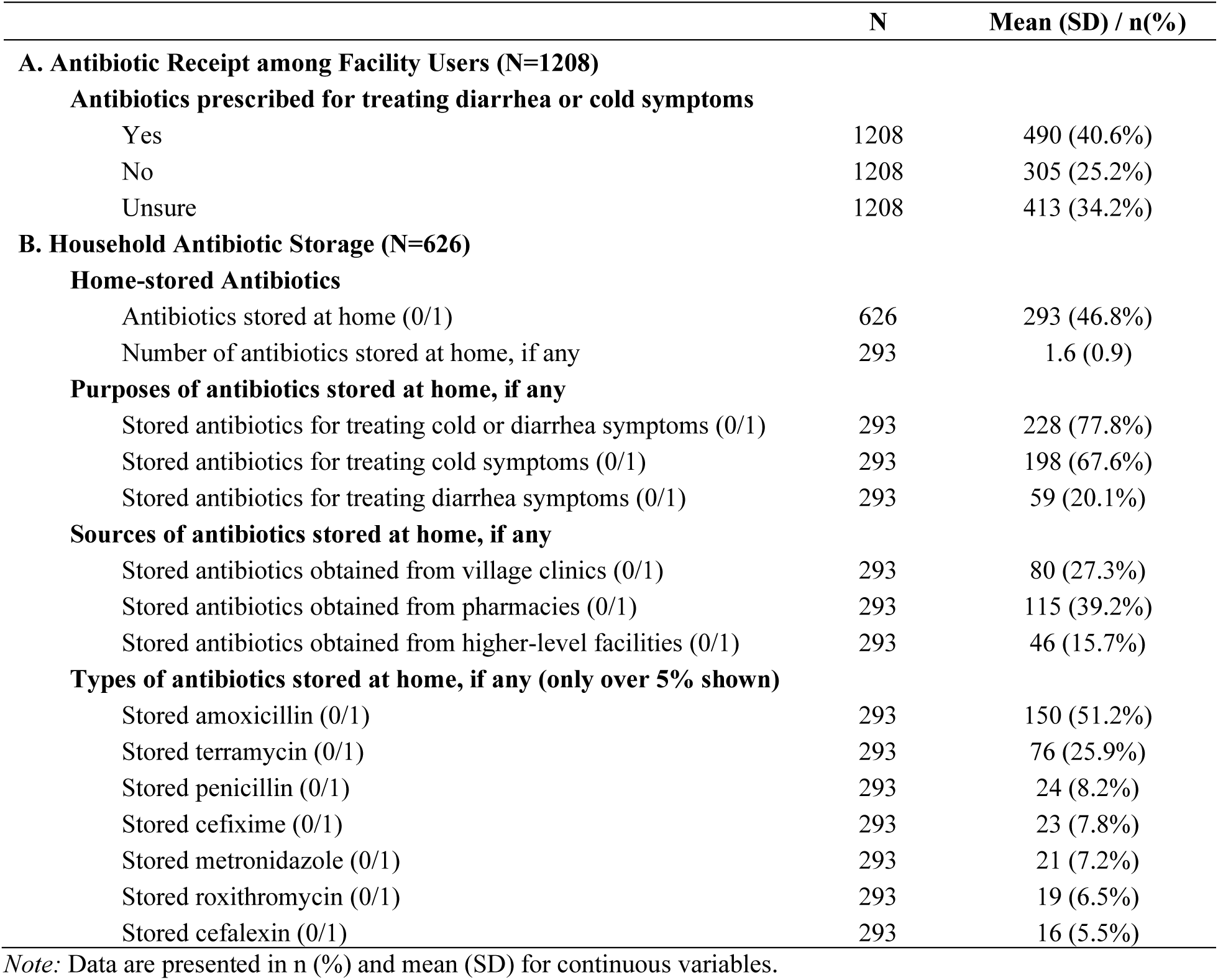
Summary statistics of household antibiotic use.

|  | N | Mean (SD) / n(%) |
|---|---|---|
| **A. Antibiotic Receipt among Facility Users (N=1208)** |  |  |
| **Antibiotics prescribed for treating diarrhea or cold symptoms** |  |  |
| Yes | 1208 | 490 (40.6%) |
| No | 1208 | 305 (25.2%) |
| Unsure | 1208 | 413 (34.2%) |
| **B. Household Antibiotic Storage (N=626)** |  |  |
| **Home-stored Antibiotics** |  |  |
| Antibiotics stored at home (0/1) | 626 | 293 (46.8%) |
| Number of antibiotics stored at home, if any | 293 | 1.6 (0.9) |
| **Purposes of antibiotics stored at home, if any** |  |  |
| Stored antibiotics for treating cold or diarrhea symptoms (0/1) | 293 | 228 (77.8%) |
| Stored antibiotics for treating cold symptoms (0/1) | 293 | 198 (67.6%) |
| Stored antibiotics for treating diarrhea symptoms (0/1) | 293 | 59 (20.1%) |
| **Sources of antibiotics stored at home, if any** |  |  |
| Stored antibiotics obtained from village clinics (0/1) | 293 | 80 (27.3%) |
| Stored antibiotics obtained from pharmacies (0/1) | 293 | 115 (39.2%) |
| Stored antibiotics obtained from higher-level facilities (0/1) | 293 | 46 (15.7%) |
| **Types of antibiotics stored at home, if any (only over 5% shown)** |  |  |
| Stored amoxicillin (0/1) | 293 | 150 (51.2%) |
| Stored terramycin (0/1) | 293 | 76 (25.9%) |
| Stored penicillin (0/1) | 293 | 24 (8.2%) |
| Stored cefixime (0/1) | 293 | 23 (7.8%) |
| Stored metronidazole (0/1) | 293 | 21 (7.2%) |
| Stored roxithromycin (0/1) | 293 | 19 (6.5%) |
| Stored cefalexin (0/1) | 293 | 16 (5.5%) |

*Note:* Data are presented in n (%) and mean (SD) for continuous variables.

Within the 626 households, 2,319 illness episodes involving cold or diarrhea symptoms were reported during the previous year for 2,261 individuals, including 1,594 episodes of cold symptoms and 725 episodes of diarrhea symptoms (Table S5). Of these episodes, 1,208 (52.1%) involved treatment-seeking at a healthcare facility, 944 (40.7%) were managed at home through self-medication, and 167 (7.2%) were untreated by self-healing. Among facility users, nearly half (48.8%) sought care at village clinics, while approximately 20% visited higher-level facilities (township, county, or city hospitals), 15.8% visited private clinics, and 10.5% sought treatment from pharmacies. These patterns are consistent with previous statistics showing that village clinics were the preferred healthcare provider when ill, followed by home self-medication.

Table 5 (Panel A) shows that, among the 1,208 illness episodes for which treatment was sought at a healthcare facility, the majority (490, 40.6%) reported receiving antibiotics, while 305 (25.2%) reported not receiving antibiotics, and 413 (34.3%) were unsure. Because this measure is observed only among healthcare facility users, it captures antibiotic exposure through formal healthcare encounters but does not reflect antibiotic use through home self-medication, an important pathway of antibiotic use in these communities. We next presented household antibiotic storage as a proxy for self-medication.

Table 5 (Panel B) shows that nearly half of the 626 households (293, 46.8%) stored antibiotics, with an average of 1.6 antibiotics stored per household. Among households storing antibiotics, most reported keeping antibiotics to treat cold or diarrhea conditions (77.8%).

Antibiotics were obtained through multiple channels: 27.3% of households stored at least one antibiotic obtained from village clinics, 39.2% from pharmacies, and 15.7% from higher-level health facilities. These source categories were not mutually exclusive. The most commonly stored antibiotics included amoxicillin (51.2%), terramycin (25.9%), penicillin (8.2%), cefixime (7.8%), metronidazole (7.2%), roxithromycin (6.5%), and cefalexin (5.5%). The storage of other antibiotics was lower than 5% and is not reported.

The household survey also assessed household knowledge regarding appropriate antibiotic use and recognition. Table S6 shows that households exhibited substantial gaps in understanding rational antibiotic use. About three-quarters of households (71.4%) believed that as long as antibiotics could treat similar conditions, they could use their families’ antibiotics, and 79.4% believed that if they had recovered from antibiotics previously, they could purchase or request the same antibiotic for similar conditions. Only 5.0% of households correctly identified that antibiotics should be taken for the full prescribed course when asked when to stop taking them, 45.2% believed that antibiotics could be discontinued once symptoms improved, and 49.8% did not know the correct answer. Households also exhibited a limited ability to identify antibiotics correctly. While approximately half recognized penicillin (46.5%) and amoxicillin (46.5%) as antibiotics, fewer correctly identified norfloxacin (24.3%), roxithromycin (25.2%), and metronidazole (23.0%). Combined, these findings suggest widespread misconceptions about antibiotic effectiveness, appropriate use, and recognition, which could lead to inappropriate antibiotic use in the home and contribute to antibiotic resistance globally.

### 5.2 Main Regression Estimates

#### 5.2.1 OLS Estimates

Table 6 (Panel A, Column 1) presents OLS estimates of the association between the clinical practice quality of village doctors and antibiotic receipt among healthcare facility users. In the primary analysis, respondents who were unsure whether antibiotics had been prescribed were treated as missing, leaving 795 healthcare facility users with complete information on antibiotic receipt. A one-standard-deviation increase in the overall clinical practice quality index of village doctors was associated with a 6.2-percentage-point reduction in the probability that patients received antibiotics for treating cold or diarrhea symptoms. The estimates were robust across alternative sets of control variables (Table S7).

**Table 6.**
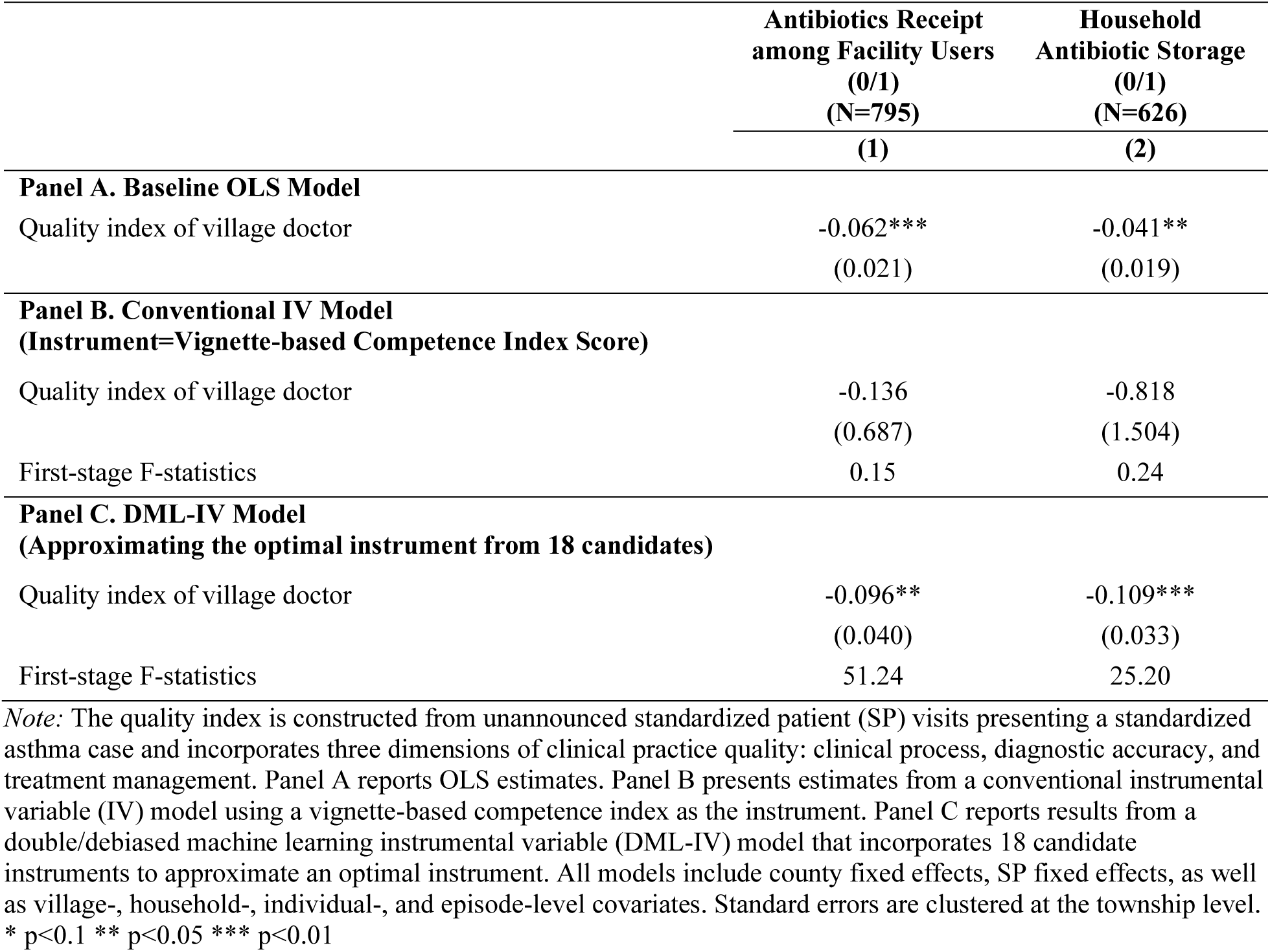
Effects of village doctor clinical practice quality on household antibiotic use.

|  | **Antibiotics Receipt among Facility Users (0/1)<br>(N=795)** | **Household Antibiotic Storage (0/1)<br>(N=626)** |
|---|---|---|
|  | **(1)** | **(2)** |
| **Panel A. Baseline OLS Model** |  |  |
| Quality index of village doctor | -0.062*** | -0.041** |
|  | (0.021) | (0.019) |
| **Panel B. Conventional IV Model<br>(Instrument=Vignette-based Competence Index Score)** |  |  |
| Quality index of village doctor | -0.136 | -0.818 |
|  | (0.687) | (1.504) |
| First-stage F-statistics | 0.15 | 0.24 |
| **Panel C. DML-IV Model<br>(Approximating the optimal instrument from 18 candidates)** |  |  |
| Quality index of village doctor | -0.096** | -0.109*** |
|  | (0.040) | (0.033) |
| First-stage F-statistics | 51.24 | 25.20 |

*Note:* The quality index is constructed from unannounced standardized patient (SP) visits presenting a standardized asthma case and incorporates three dimensions of clinical practice quality: clinical process, diagnostic accuracy, and treatment management. Panel A reports OLS estimates. Panel B presents estimates from a conventional instrumental variable (IV) model using a vignette-based competence index as the instrument. Panel C reports results from a double/debiased machine learning instrumental variable (DML-IV) model that incorporates 18 candidate instruments to approximate an optimal instrument. All models include county fixed effects, SP fixed effects, as well as village-, household-, individual-, and episode-level covariates. Standard errors are clustered at the township level.
\* p<0.1 \*\* p<0.05 \*\*\* p<0.01

Since a substantial share of facility users in our study were unsure whether they had received antibiotics, we performed sensitivity analyses using multiple imputations by chained equations (MICE) to impute missing antibiotic-use responses based on rich individual-, household-, and community-level data (Azur et al., 2011). This procedure iteratively imputes missing values based on observed characteristics for a given individual and on relationships with other variables in the sample for similar individuals, and repeats this process multiple times. Unlike simple approaches such as mean imputation, which treat imputed values as if they were known true values, multiple imputation explicitly recognizes and quantifies the uncertainty around missing data by generating multiple predictions for each missing observation and creating multiple complete datasets. Following this procedure, we generated five imputed datasets and estimated models separately in each imputed dataset. Estimates were combined using Rubin’s rules, accounting for the variability within each imputed dataset and the uncertainty arising from the imputation process. Results from the imputed datasets were highly consistent with those from the complete-case analysis (Table S8).

We note that the primary analysis included all patients who sought formal healthcare rather than restricting the sample to village-clinic users. Village doctor quality may influence not only prescribing during village-clinic encounters but also healthcare-seeking decisions. Restricting the analysis to village-clinic users would therefore condition on a potentially endogenous healthcare-seeking decision and would estimate effects only among a selected subgroup. Our primary specification instead captures the broader community-level effect of local primary care quality on antibiotic receipt during formal healthcare encounters, integrating any effects operating through provider choices.

We also report estimates restricted to village-clinic users as a sensitivity analysis (Table S9). When restricting the analysis to village-clinic users, the estimates remained similar in magnitude (−5.5 vs. −6.2 percentage points), although statistical significance declined to the 10% level. The confidence interval widened substantially because the restricted sample was reduced substantially compared to the size of the primary analysis. These results are generally consistent with the primary results but provide less precise evidence regarding antibiotic receipt specifically during village clinic encounters.

Table 6 (Panel B, Column 2) presents OLS estimates for household antibiotic storage. Unlike antibiotic receipt during healthcare encounters, this outcome was directly measured by trained enumerators during household medicine inventory assessment and was available for all households. A one-standard-deviation increase in the clinical practice index was associated with a 4.1-percentage-point probability that households living in the same village stored antibiotics at home. The results were robust across alternative sets of control variables (Table S10).

#### 5.2.2 DML-IV Estimates

We first present results from a conventional IV model to illustrate the limitations of the conventional IV model in this setting (Table 6, Panel B). Because it is impractical to include a large number of instruments in a conventional IV specification, we estimated a conventional IV model using a single instrument, the asthma vignette-based competence index. The asthma vignette followed protocols similar to those used in SP visits. However, the vignette captured clinical diagnoses and treatment in hypothetical scenarios, whereas the unannounced SP visits measured their actual clinical practice under real-world conditions.

Table 6 (Panel B) suggests that, although using comparable instruments and protocols, the vignette-based competence poorly predicted the clinical practice quality of village doctors as measured by unannounced SP visits. The first-stage regression yielded very low F-statistics (0.15-0.24), indicating a severe weak-instrument problem. These results also suggest that provider knowledge alone does not fully determine real-world clinical practice, highlighting the role of other factors in shaping provider behavior during actual patient encounters. Accordingly, the conventional IV estimates are not interpreted further because weak instruments can produce biased and unstable estimates.

Table 6 (Panel C) reports estimates from the DML-IV model, which builds on the previous specification but combines 18 candidate instruments to approximate an optimal instrument, accommodating flexible modeling of high-dimensional instruments and controls. The resulting optimal instrument exhibited strong first-stage predictive power, with F-statistics of 51.24 for antibiotic receipt among facility users and 25.20 for household antibiotic storage.

The DML-IV estimates indicate that the higher clinical practice quality of village doctors contributed to significantly lower household antibiotic use. A one-standard deviation increase in the clinical practice quality index of village doctors led to reductions in the probability that facility users received antibiotics for treating cold or diarrhea symptoms by 9.6 percentage points and the likelihood that households living in the same communities stored antibiotics at home by 10.9 percentage points. To contextualize these magnitudes, relative to baseline rates of 61.6% antibiotic receipt among healthcare facility users and 46.8% household antibiotic storage, these estimates correspond to reductions of 15.6% and 23.3% in antibiotic use, respectively.

Overall, both the OLS and the DML-IV estimates suggest that higher clinical practice quality among village doctors was associated with lower antibiotic receipt through formal healthcare encounters and home antibiotic storage for future self-medication. The DML-IV estimates were larger in magnitude than the OLS estimates. In addition to correcting for potential bias in the OLS specification, the DML-IV estimates may also reflect a local average treatment effect (LATE)-type interpretation, capturing the effect of improvements in provider quality along the particular margin shifted by the instruments used in this study. In our setting, these instruments primarily reflect upstream provider attributes, such as training, credentials, and latent clinical competence, which plausibly translate into improvement in real-world clinical practice. To the extent that these upstream characteristics improve actual clinical practice, the larger DML-IV estimates suggest that quality improvements operating through these channels may have particularly stronger effects on household antibiotic use.

#### 5.2.3 Plausibly Exogeneous IV

Although the DML-constructed instrument exhibited strong first-stage predictive performance for village doctors’ clinical practice quality, the causal interpretation of the IV estimates remains conditional on the exclusion restriction. To assess the sensitivity of the IV estimates to potential violations of the exclusion restriction, we employed the plausibly exogenous IV bounding approach proposed by Conley et al. (2012), using the union of confidence intervals (UCI) method, following recent applications (Barschkett et al., 2026; Nguyen-Phung et al., 2026). Under the conventional exclusion restriction, the constructed instrument is assumed to affect household antibiotic use only though village doctors’ clinical practice quality, implying no direct effect of the instrument on the outcomes (*δ* = 0). The plausibly exogenous approach relaxes this assumption by allowing small, nonzero direct effects (*δ* ≈ 0) and examining how inference about the main coefficient changes under varying degrees of relaxation of the exclusion restriction.

Figure 2 illustrates how the IV estimates and their 95% confidence intervals vary as the assumed direct effect of the constructed instrument (*δ*) ranges from −0.1 to 0.1. The dash lines represent the lower and upper confidence limits under different values of direct effects (*δ*). The results suggest that the confidence intervals remain below zero when *δ* takes positive values, whereas sufficiently negative values of *δ* could cause the intervals to include zero.

**Figure 2.**
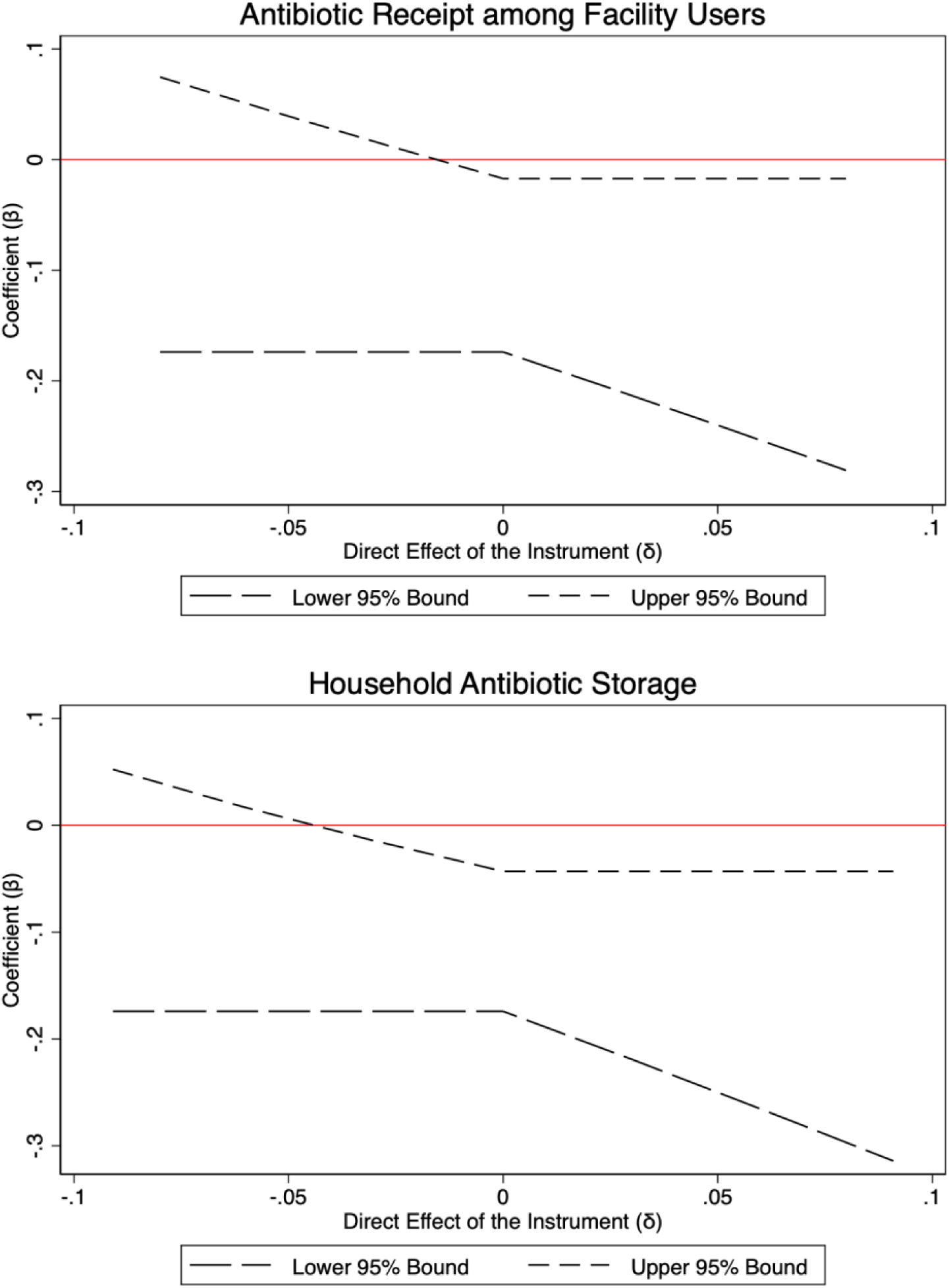
Sensitivity of DML-IV Estimates to Violations of the Exclusion Restriction. *Note.* The figure presents the results of the plausibly exogenous instrumental variables sensitivity analysis following Conley et al. (2012). The analysis uses cross-fitted, residualized variables derived from the double/debiased machine learning instrumental variables (DML-IV) estimation. The horizontal axis represents the assumed direct effect of the constructed instrument on each outcome (δ), ranging from −0.1 to 0.1, with δ = 0 corresponding to the conventional exclusion restriction. The vertical axis represents the coefficient on provider clinical practice quality (β). The long-dashed and short-dashed lines represent the lower and upper bounds of the 95% confidence intervals under each assumed value of δ.

We next used the cross-fitted, residualized variables from the DML-IV estimation to estimate the reduced-form relationship between the constructed instrument and each outcome. The estimated reduced-form coefficients were −0.0799 for antibiotic receipt among facility users and −0.0909 for household antibiotic storage. Table S11 reports that the 95% UCI as we progressively relax the exclusion restriction, allowing the maximum absolute direct effect of the instrument to range from 0% to 100% of the absolute reduced-form effect, with the permitted direct effects bounded symmetrically around zero. At 0%, the conventional exclusion restriction is imposed, assuming no direct effect of the instrument on the outcomes. At 10%, for example, the permitted direct effect is bounded between −10% and 10% of the absolute reduced-form magnitude. The results show that the 95% UCI first includes zero when the maximum permitted direct effect reaches 20% of the reduced-form magnitude for antibiotic receipt and 50% for household antibiotic storage.

Overall, the plausibly exogenous IV analyses indicate that the negative IV estimates remain statistically distinguishable from zero under certain relaxations of the exclusion restriction, although their sensitivity varies across outcomes. The IV estimate for household antibiotic storage exhibits greater robustness to potential violations of the exclusion restriction than the estimate for antibiotic receipt during healthcare encounters.

### 5.3 Exploratory Analysis by Clinical Practice Quality Domain

We next conducted exploratory analyses to examine how different domains of village doctors’ clinical practice quality (clinical process, diagnostic accuracy, and treatment management) were associated with household antibiotic use. All exploratory analyses were estimated using OLS models.

Figure 3 presents OLS estimates of the association between the three clinical practice quality domains and household antibiotic use. When decomposing the overall quality index into process quality, diagnostic quality, and treatment management quality, we found that process quality and diagnostic quality, which capture general clinical competencies in adequate assessment and diagnostic reasoning, were most strongly associated with lower antibiotic receipt among healthcare facility users for treating cold or diarrhea symptoms. A one-standard deviation increase in process quality and diagnostic quality was associated with reductions of 6.9 percentage points and 5.2 percentage points, respectively, in the probability that individuals seeking care for cold or diarrhea symptoms received antibiotics. By contrast, treatment management quality, which captures the appropriateness of disease management and medication recommendations, was more strongly associated with household antibiotic storage, corresponding to a 5.6-percentage-point reduction. Diagnostic quality was also associated with a 3.7-percentage-point reduction in the likelihood that households stored antibiotics, whereas the association with process quality was only marginally statistically significant.

**Figure 3.**
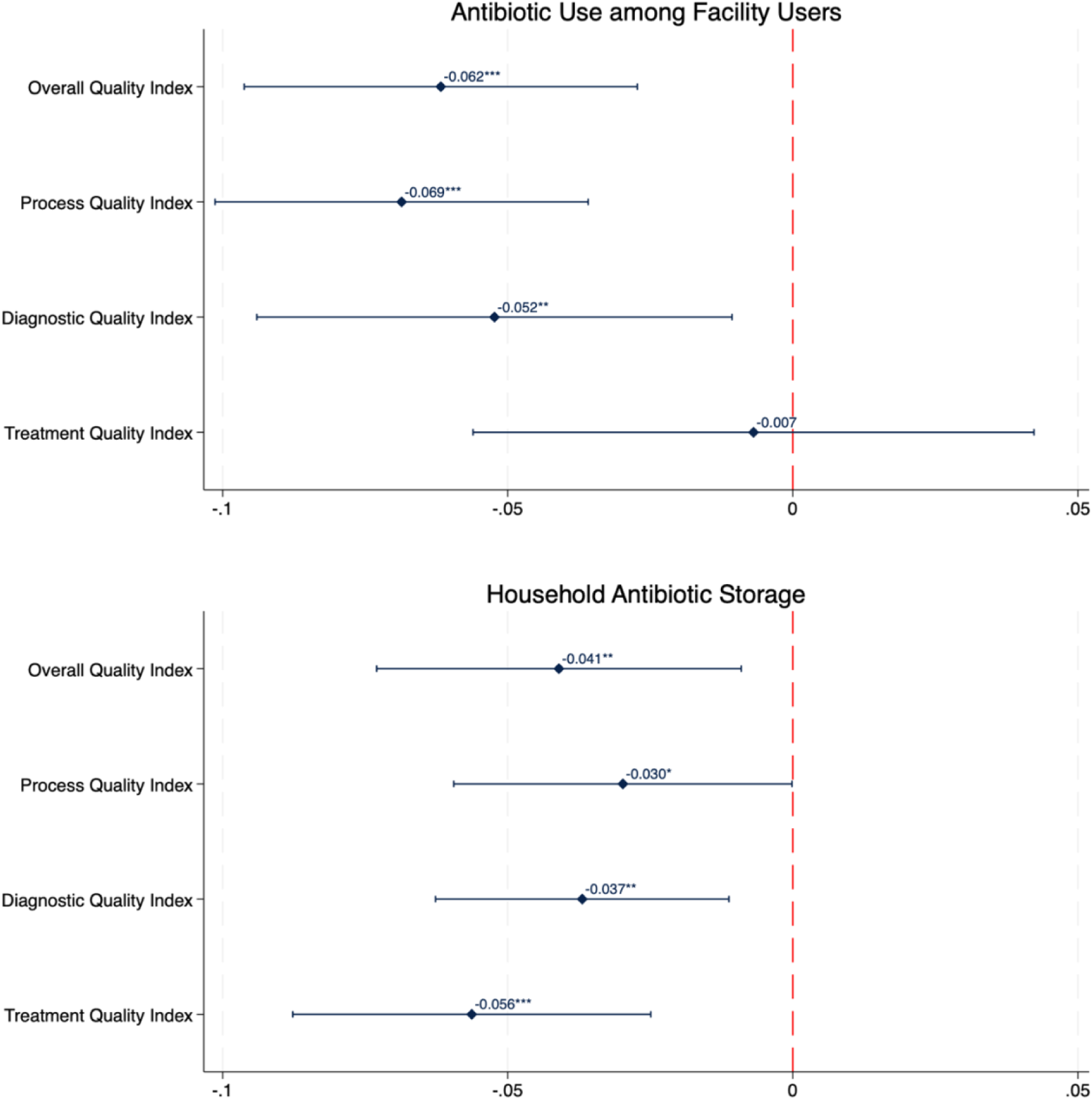
OLS estimates for quality domains and household antibiotic use. *Note:* This figure plots the OLS coefficient estimates for the associations between different domains of village doctor clinical practice quality (clinical process, diagnostic accuracy, and treatment management) and two measures of household antibiotic use. All models include county fixed effects, standardized patient (SP) fixed effects, as well as village-, household-, individual-, and episode-level covariates. Standard errors are clustered at the township level. * p<0.1 ** p<0.05 *** p<0.01

These findings suggest that different domains of provider quality may shape different aspects of household antibiotic use. More systematic clinical assessment and diagnostic reasoning may primarily shape the decision of whether antibiotics are ultimately prescribed. By contrast, better treatment management quality may reduce the quantity of unnecessary antibiotics entering households. Combining with earlier descriptive evidence that most household-stored antibiotics were sourced from village clinics and pharmacies, these results may suggest that medication recommendations may be more observable to patients, potentially shaping household accumulation of antibiotics available for later use, or even potentially shaping household medication-seeking behaviors for similar illness episodes.

### 5.4 Exploratory Analysis of Potential Pathways

To explore potential pathways underlying the main estimates, we examined exploratory intermediate outcomes measured at both the illness-episode and household levels, corresponding to the margins outlined in our conceptual framework. At the illness-episode level, we examined village clinic utilization as an extensive-margin outcome and antibiotic receipt conditional on visiting a village clinic as an intensive-margin outcome. At the household level, we examined the relationship between objectively measured clinical quality, household perceptions of village clinic quality, and stated preferences for village clinic care as potential factors underlying extensive-margin responses. We also assessed antibiotic knowledge and recognition as potential downstream pathways related to household antibiotic storage and self-medication. These analyses are exploratory and were estimated using OLS models.

For the illness-episode-level analyses, we found little evidence that village doctor clinical practice quality substantially affected village clinic utilization, suggesting limited changes along the extensive margin (Table S12). In contrast, conditional on visiting a village clinic, higher village doctor clinical practice quality was associated with a significantly lower probability of receiving antibiotics, suggesting that provider quality may influence antibiotic use primarily along the intensive margin, through improvements in clinical decision-making and prescribing during healthcare encounters, rather than through substantial changes in village clinic utilization.

At the household level, we further explored factors that may help explain the limited responses along the extensive margin. We found limited association between objectively measured clinical practice quality and household perceived quality of village clinics. In contrast, household perceived quality of village clinics was strongly associated with their stated preferences for village clinic care (Table S13). These patterns are consistent with a potential disconnect between objective and perceived provider quality: households may respond to their perceptions of provider quality when making healthcare choices, while these perceptions may not closely reflect the clinical practice quality captured by standardized patient assessments.

We also examined household-level pathways potentially related to downstream antibiotic storage and self-medication. We combined household responses to multiple questions on appropriate antibiotic use and recognition into composite indices but found little evidence that village doctor clinical practice quality was associated with either measure (Table S14). Thus, the observed reductions in household antibiotic storage does not appear to be explained by measurable changes in household antibiotic knowledge or recognition. Instead, the stronger association between village doctor treatment management quality and household antibiotic storage, together with the observation that home-stocked antibiotics were commonly sourced from village clinics and pharmacies, may suggest a potential downstream consequence of prescribing and dispensing during healthcare encounters. First, more appropriate medication prescribing may reduce the quantity of unnecessary antibiotics entering households, thereby limiting the accumulation of leftover medications available for future self-medication. Second, repeated exposure to medication recommendations may gradually shape households’ expectations and future medication-seeking behaviors for similar illness episodes. Nevertheless, these hypotheses warrant further investigation.

## 6. Discussion

This paper examines how frontline primary care quality shapes community antibiotic use, focusing on a low-resource setting where access to quality healthcare remains limited while access to antibiotics is relatively unrestricted. We draw on unique community-based data from remote communities in rural China, linking objective measures of village doctors’ clinical practice quality, measured through unannounced standardized patient clinical visits, to household-level antibiotic use data collected in the same communities. To mitigate potential endogeneity concerns, we employed an IV approach integrated within a DML framework to address a many-weak-instruments problem. In our setting, conventional IV specifications were constrained by multiple weak instruments and limited first-stage predictive power. We leveraged extensive provider-level information to construct candidate instruments for predicting their clinical practice quality. We employed a flexible DML-IV framework to aggregate predictive information across multiple individually weak but jointly informative instruments using an ensemble of machine learning algorithms to approximate an optimal instrument. This approach strengthened first-stage predictive performance while retaining the interpretation of conventional IV estimation, although causal interpretation remains conditional on the underlying identifying assumptions. More broadly, our study illustrates how causal machine learning can complement traditional econometric methods in global health economics research by leveraging rich, multidimensional data to address estimation challenges in complex high-dimensional settings.

Our findings show that, despite substantial limitations in healthcare quality, village providers remained the primary source of care for most families living in these communities. Antibiotic receipt for treating cold or diarrhea symptoms was common among facility users during healthcare encounters, and approximately half of households stored antibiotics at home, primarily for treating these conditions. Both the OLS and DML-IV analyses suggest that village doctor clinical practice quality was associated with reductions in inappropriate antibiotic use during healthcare encounters and household storage of antibiotics.

Our exploratory analyses suggest that different dimensions of clinical practice quality may matter along different aspects of antibiotic use. Process quality, particularly systematic history-taking and clinical examination, appears to play a more prominent role in reducing inappropriate antibiotic prescribing during patient encounters. Providers, who more consistently ask relevant questions and perform appropriate assessments, may be better able to distinguish self-limiting conditions from bacterial infections, thereby reducing precautionary antibiotic prescribing when diagnostic confidence is low. By contrast, treatment management quality, particularly how medications are ultimately prescribed, may be more observable to patients and thus more influential in shaping household storage of antibiotics, generating spillover effects on antibiotic use beyond formal clinical encounters. Treatment decisions and the medicines patients take home may be highly visible and memorable, shaping how families store and self-medicate with antibiotics long after the clinical visit. Most importantly, diagnostic quality appears to matter for both outcomes, suggesting that village doctors’ ability to identify illness accurately may be central to reducing overall antibiotic use in community settings.

We also explored several potential pathways corresponding to the extensive and intensive margins outlined in our conceptual framework. Along the extensive margin, we found little evidence that provider quality substantially changed household healthcare-seeking behavior or the use of village clinics. In contrast, along the intensive margin, conditional on visiting a village clinic, higher provider quality was associated with a lower probability of patients receiving antibiotics. We also found little evidence that provider quality substantially changed household antibiotic knowledge or recognition. Together, these exploratory analyses suggest that reductions in community antibiotic use may be more likely to operate through improvements in clinical decision-making and antibiotic prescribing during formal healthcare encounters than through changes in care-seeking behavior, with potentially important downstream consequences for household antibiotic storage and self-medication.

In our data, home-stocked antibiotics were mainly sourced from village clinics (27%) and pharmacies (39%), suggesting that antibiotics entering households through village clinic encounters and pharmacy purchases may contribute importantly to household antibiotic storage. Additionally, 79.4% of households believed that if they had previously recovered from antibiotics, they could purchase or request the same antibiotic for similar conditions in the future. Together, these findings suggest that inappropriate antibiotic prescribing during healthcare encounters may not only contribute to the household accumulation of leftover antibiotics available for future self-medication, but also potentially shape households’ future medication-seeking behaviors when similar symptoms arise.

The findings of this study should be interpreted in light of several limitations. First, antibiotic receipt during healthcare encounters was based on household self-report and may therefore be subject to recall error and misclassification. However, tracing antibiotic use for every healthcare encounter was logistically difficult and prohibitively costly in this setting, making household reporting the most feasible approach. Additionally, information on antibiotic receipt during healthcare encounters was missing for a substantial proportion of facility users. To assess the sensitivity of our findings to missing data, we used multiple imputations by chained equations to generate imputed datasets. We acknowledge that this approach assumes that missingness depends on observed variables, although the availability of rich, multi-level data on communities, providers, and households helps support the plausibility of this assumption. We also complemented the self-reported outcome with an objective measure of household antibiotic storage based on in-person medicine inventory audits conducted by trained enumerators. The consistency of results across these two distinct measures provides reassurance that our main findings are unlikely to be driven by reporting bias.

Second, although the DML framework strengthens first-stage prediction in the presence of many weak but jointly informative instruments, it does not relax the fundamental identifying assumptions of the IV approach. The exclusion restriction remains inherently untestable and must be justified based on institutional knowledge and economic reasoning. To support the plausibility of this assumption, we selected provider characteristics with conceptually defensible exclusion restrictions, documented the long-term stability of village doctor placement, and adjusted for possible observable factors capturing plausible alternative pathways. We additionally employed plausibly exogenous IV bounding analyses and showed that the negative IV estimates remained statistically distinguishable from zero under certain relaxations of the exclusion restriction. Nevertheless, potential violations of the identifying assumptions through unobserved factors and alternative pathways cannot be ruled out, warranting caution in causal interpretations.

Inappropriate antibiotic use remains one of the most pressing global public health challenges. Evidence from this study suggests that strengthening the clinical practice quality of frontline providers in resource-constrained settings may help address inappropriate community antibiotic use, both during healthcare encounters and through its potential downstream implications for household antibiotic storage and subsequent self-medication. Improving frontline primary care quality, whether through capacity-building, strengthened supervision, or emerging clinical decision support and diagnostic technologies, may therefore represent an integral component of antibiotic stewardship in LMICs, complementing regulatory approaches that primarily focus on restricting antibiotic access.

## Data Availability

All data produced in the present study are available upon reasonable request to the authors.

## Supplementary Materials

### A. Appendix Figures

**Figure S1.**
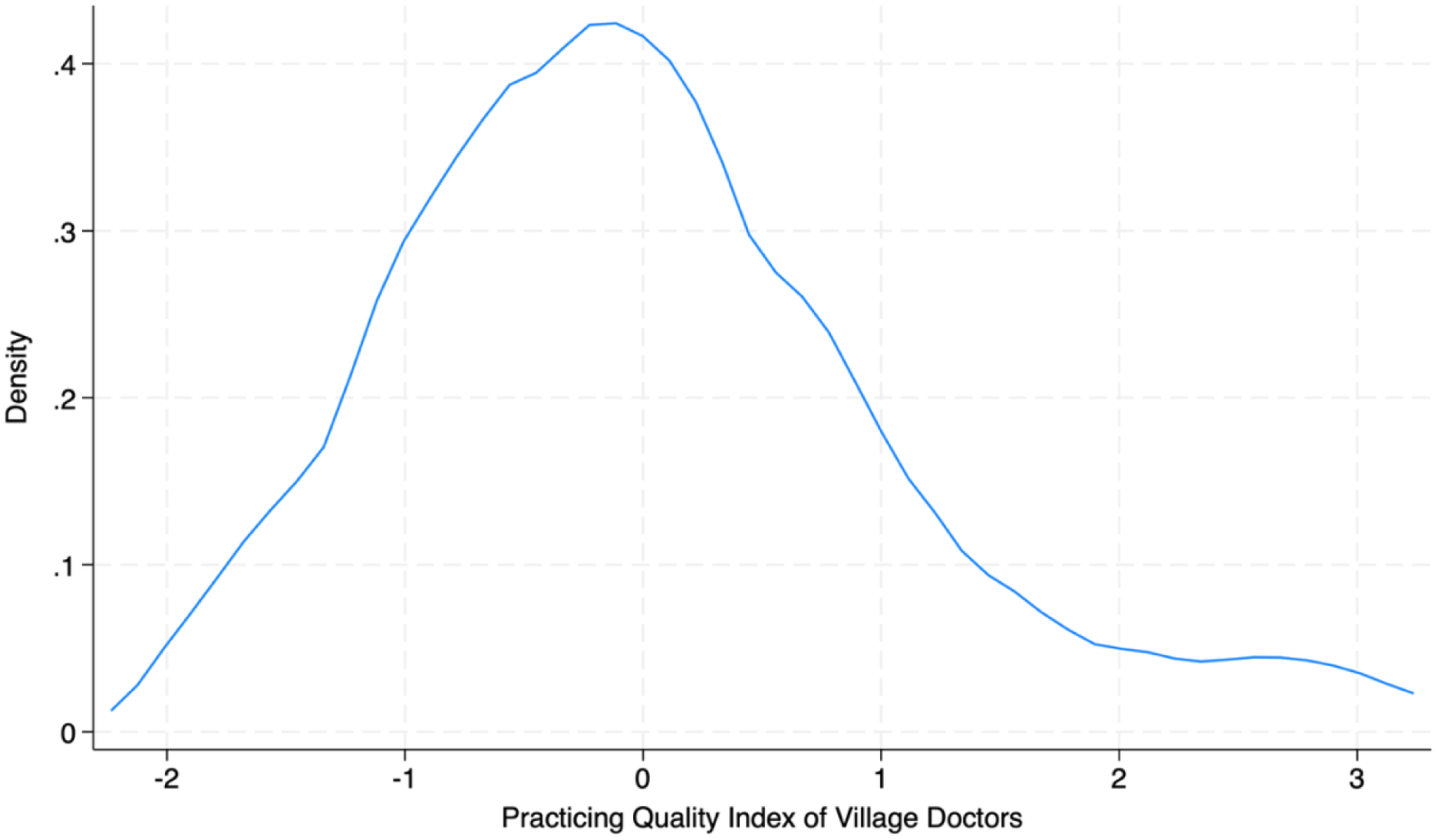
Distribution of the constructed quality index of village doctors. *Note:* This figure shows that the constructed quality index follows a normal distribution, illustrating that the constructed measure captures meaningful variations in the clinical practice quality across village doctors. The quality index was constructed for each village doctor based on data collected from unannounced standardized patient clinical visits, with higher values representing higher quality practice.

**Figure S2.**
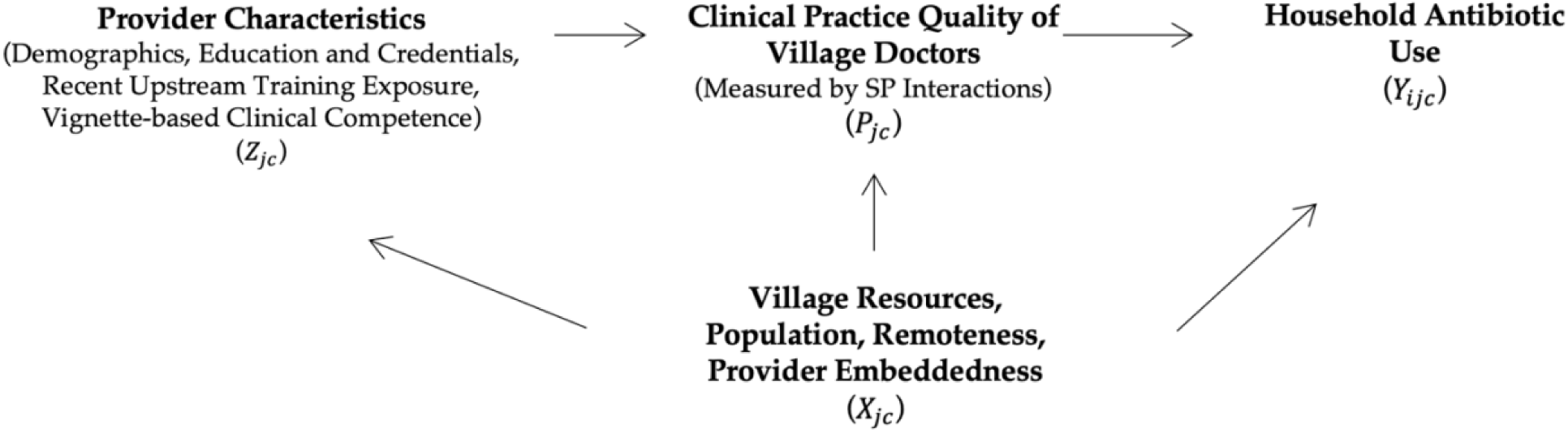
Instrumental variable (IV) approach framework.

**Figure S3.**
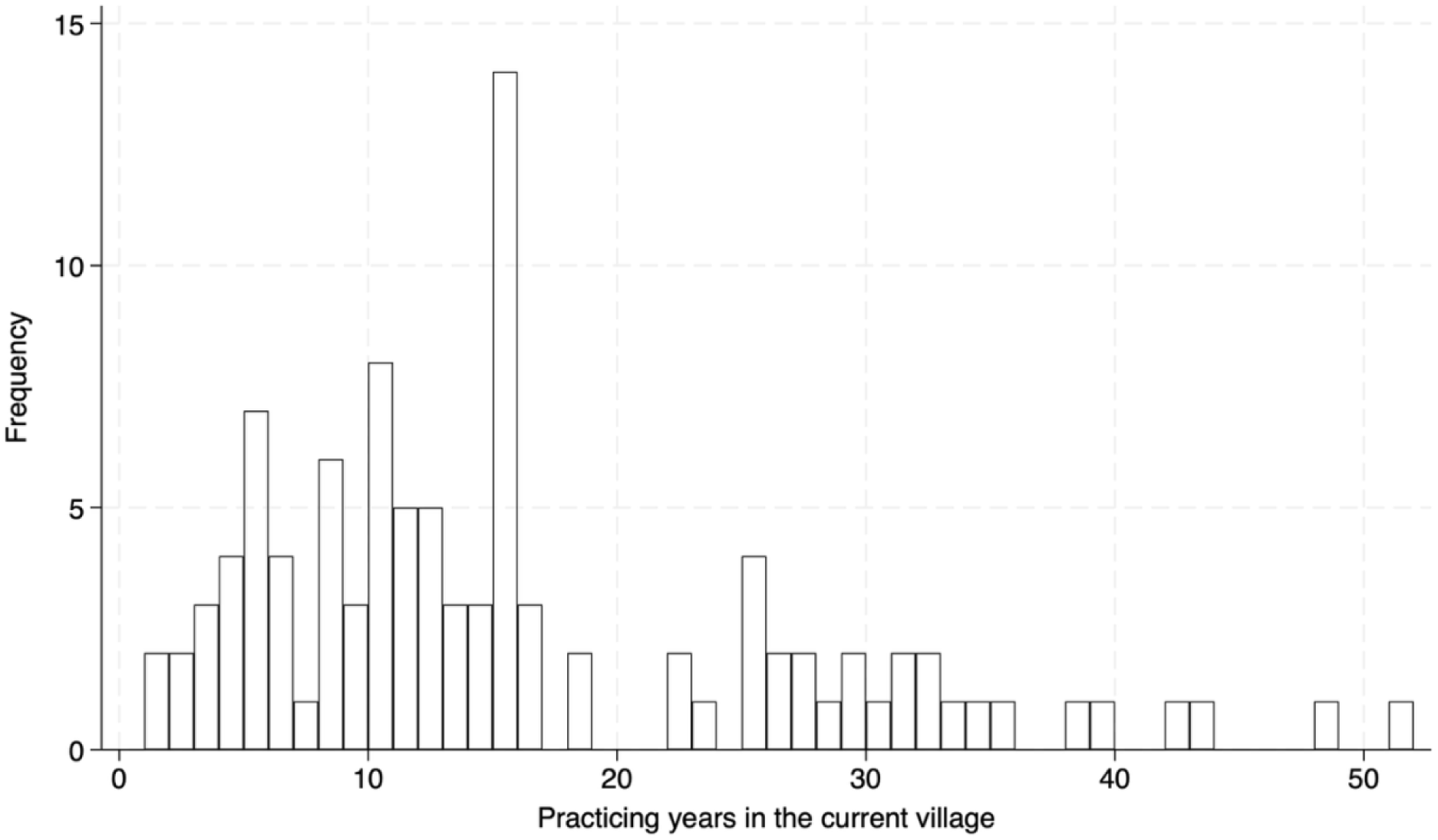
Distribution of the practicing years of village doctors in their current village.

**Figure S4.**
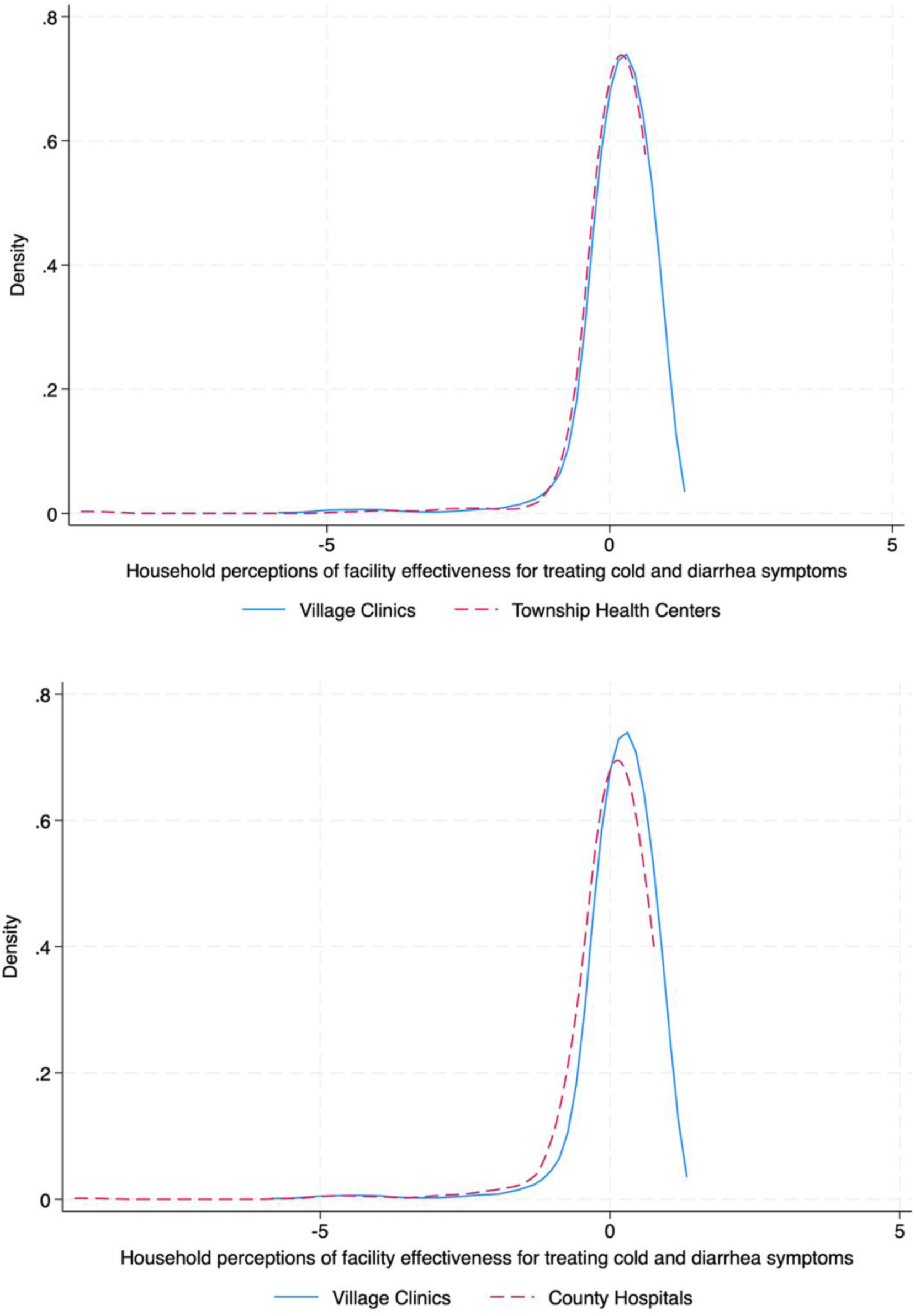
Household perceptions of facility effectiveness for cold or diarrhea symptoms.

**Figure S5.**
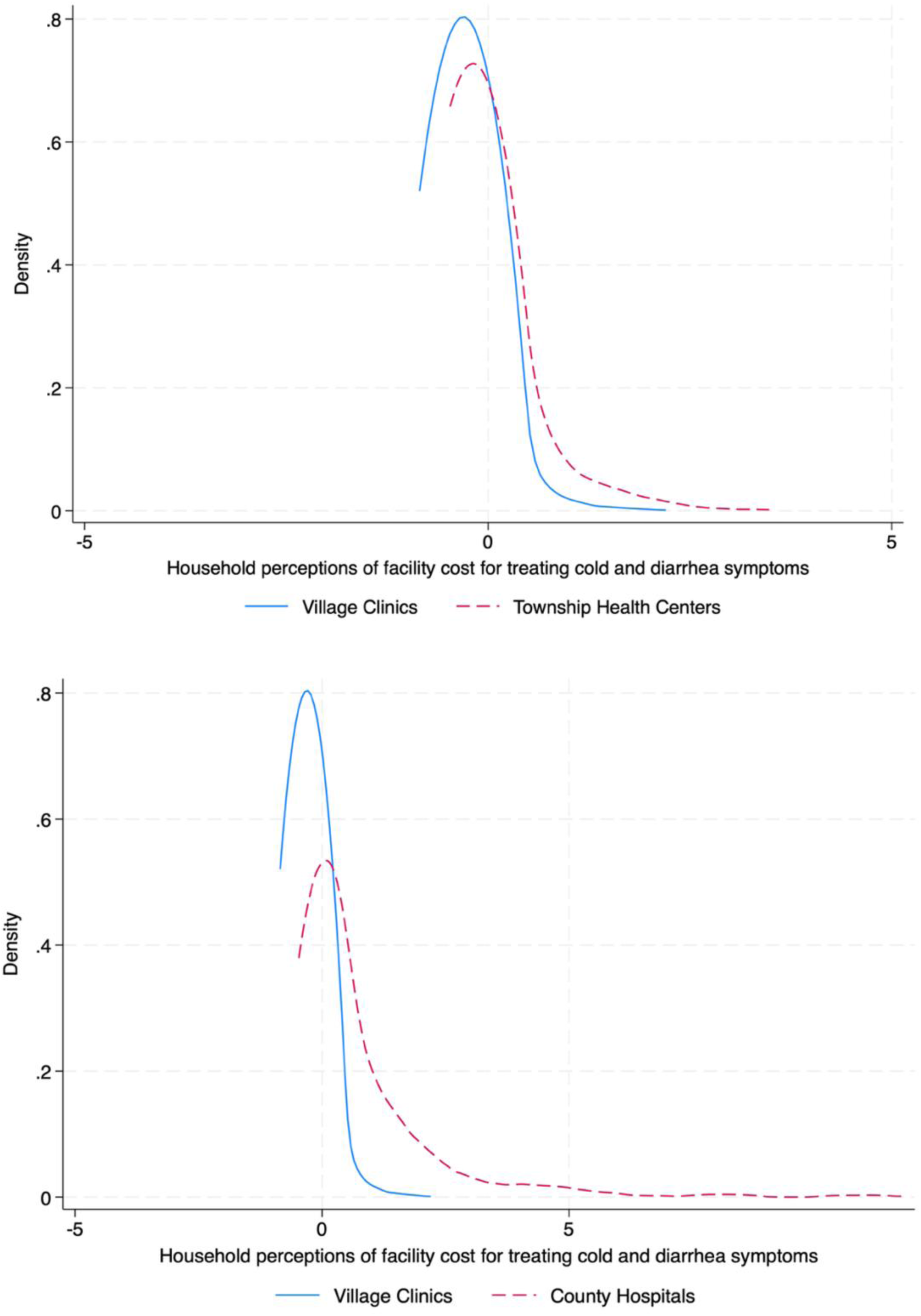
Household perceptions of facility cost for treating cold or diarrhea symptoms.

### B. Appendix Tables

**Table S1.** List of potential instruments for predicting provider clinical practices.

| **Domains** | **No** | **Variables** |
|---|---|---|
| Demographics | 1 | Age |
|  | 2 | Gender |
|  | 3 | Ethnicity |
| Credentials | 4 | Medical practice years |
|  | 5 | Education |
|  | 6 | Formal medical diploma |
|  | 7 | Medical practice license |
| Recent training exposure | 8 | Township-level medical training last year |
|  | 9 | County-level training last year |
|  | 10 | City-level training last year |
|  | 11 | Remote online training last year |
| Clinical vignette performance | 12 | Overall competence index score |
|  | 13 | Asthma vignette index score |
|  | 14 | Diarrhea vignette index score |
|  | 15 | Cold vignette index score |
|  | 16 | Clinical process competence index score |
|  | 17 | Diagnostic competence index score |
|  | 18 | Treatment competence index score |

**Table S2.** Competence measure of village doctors (asthma case, N=103).

|  | **n (%)** |
|---|---|
| **A. Recommended Questions for Diagnosing Asthma** |  |
| 1. Symptoms onset last time | 0 (0.0%) |
| 2. The progression of the disease | 9 (8.7%) |
| 3. Approaches to alleviating symptoms | 6 (5.8%) |
| 4. Triggers of symptoms | 20 (19.4%) |
| 5. Intensity and duration of symptoms | 10 (9.7%) |
| 6. Difficulty breathing | 12 (11.7%) |
| 7. Onset of first symptoms | 52 (50.5%) |
| 8. Sounds of breathing (wheezing) | 1 (1.0%) |
| 9. Cold/fever | 30 (29.1%) |
| 10. Coughing/expectoration | 61 (59.2%) |
| 11. Medical history of the family | 5 (4.9%) |
| 12. Other diseases | 37 (35.9%) |
| 13. History of childhood illnesses | 1 (1.0%) |
| **B. Recommended Exams for Diagnosing Asthma** |  |
| 1. Chest/back auscultation | 17 (16.5%) |
| 2. Pulmonary ventilation test | 5 (4.9%) |
| 3. Bronchodilator test (airway reversible test) | 1 (1.0%) |
| 4. Physical examination | 28 (27.2%) |
| 5. Chest X-ray test | 24 (23.3%) |
| 6. Routine blood tests | 13 (12.6%) |
| 7. Unvoiced double lung percussion | 0 (0.0%) |
| **C. Diagnosis** |  |
| Gave correct diagnosis | 31 (30.1%) |
| Gave correct or partially correct diagnosis | 31 (30.1%) |
| **D. Treatment** |  |
| Correct medication: inhaled/oral corticosteroids and beta-receptor agonists | 15 (14.6%) |
| Did not give antibiotics | 71 (68.9%) |

*Note:* Data are presented in n (%). Correct diagnoses are those directly related to asthma or bronchial asthma. Partially correct diagnoses are those related to dyspnea but not specifically related to asthma.

**Table S3.** Competence measure of village doctors (cold case, N=103).

|  | n (%) |
|---|---|
| **A. Recommended Questions for Diagnosing Cold** |  |
| 1. Symptom onset | 35 (34.0%) |
| 2. Fever | 30 (29.1%) |
| 3. Cough | 38 (36.9%) |
| 4. Characteristics and color of sputum | 34 (33.0%) |
| 5. Itching or pain in the throat | 43 (41.7%) |
| 6. Color and characteristics of the snivel | 29 (28.2%) |
| 7. Fatigue | 26 (25.2%) |
| 8. Headache | 17 (16.5%) |
| 9. Chest pain | 7 (6.8%) |
| 10. Shortness of breath | 3 (2.9%) |
| 11. Similar symptoms in surroundings | 0 (0.0%) |
| 12. Treatment and/or medication history | 23 (22.3%) |
| **B. Recommended Exams for Diagnosing Cold** |  |
| 1. Body temperature | 67 (65.0%) |
| 2. Checking the throat | 6 (5.8%) |
| 3. Routine blood tests | 16 (15.5%) |
| 4. Checking lymph nodes | 0 (0.0%) |
| **C. Diagnosis** |  |
| Gave correct diagnosis | 7 (6.8%) |
| Gave correct or partially correct diagnosis | 77 (74.8%) |
| **D. Treatment** |  |
| Lifestyle recommendations and/or advised medications for symptomatic reliefs | 54 (52.4%) |
| Correct advice for antibiotic use | 62 (60.2%) |

*Note:* Data are presented in n (%). Two versions of cold cases were provided, one viral cold and one bacterial cold. Correct diagnoses are those directly related to viral/bacterial colds or viral/bacterial upper respiratory tract infections. Partially correct diagnoses are those related to colds but not specifically related to viral/bacterial colds. The correct advice for using antibiotics refers to prescribing antibiotics for bacterial colds and no antibiotics for viral colds.

**Table S4.** Competence measure of village doctors (diarrhea case, N=103).

|  | n (%) |
|---|---|
| **A. Recommended Questions for Diagnosing Diarrhea** |  |
| 1. Characteristics of the stool | 36 (35.0%) |
| 2. Frequency of stools | 69 (67.0%) |
| 3. Blood or mucus in the stool | 46 (44.7%) |
| 4. The amount of stool | 5 (4.9%) |
| 5. Characteristics of urine (including color) | 6 (5.8%) |
| 6. Urination last time | 0 (0.0%) |
| 7. Fever | 34 (33.0%) |
| 8. Smelly stools | 4 (3.9%) |
| 9. Worms in the stool | 0 (0.0%) |
| 10. Abdominal pain | 34 (33.0%) |
| 11. Vomiting | 44 (42.7%) |
| 12. Diet | 59 (57.3%) |
| 13. Weaknesses | 8 (7.8%) |
| 14. Hygiene habits (especially hand washing) | 0 (0.0%) |
| 15. Request that the child be brought to the clinic | 0 (0.0%) |
| 16. Similar symptoms in surroundings | 0 (0.0%) |
| **B. Recommended Exams for Diagnosing Diarrhea** |  |
| 1. Stool examination | 21 (20.4%) |
| 2. Mucosal humidity examination (e.g., lips etc.) | 1 (1.0%) |
| 3. Abdominal palpation | 14 (13.6%) |
| 4. Body temperature | 24 (23.3%) |
| 5. Weight | 7 (6.8%) |
| **C. Diagnosis** |  |
| Gave correct diagnosis | 3 (2.9%) |
| Gave correct or partially correct diagnosis | 66 (64.1%) |
| **D. Treatment** |  |
| Gave rehydration salts | 22 (21.4%) |
| Gave montmorillonite | 25 (24.3%) |
| Gave probiotics | 11 (10.7%) |
| Gave no antidiarrheals (other than montmorillonite) | 101 (98.1%) |
| Correct advice for antibiotic use | 37 (35.9%) |

*Note:* Data are presented in n (%). Two versions of diarrhea cases were provided, one viral diarrhea and one bacterial diarrhea. Correct diagnoses are those directly related to viral or bacterial diarrhea. Partially correct diagnoses are those related to diarrhea or enteritis but not specifically related to viral/bacterial diarrhea. The correct advice for using antibiotics refers to prescribing antibiotics for bacterial diarrhea and no antibiotics for viral diarrhea.

**Table S5.** Healthcare-seeking behavior and antibiotic use for cold or diarrhea symptoms.

|  | All Episodes | Cold Episodes | Diarrhea Episodes |
|---|---|---|---|
| **Illness episodes** | **2319** | **1594** | **725** |
| Self-healing (Do nothing) | 167 (7.2%) | 99 (6.2%) | 68 (9.4%) |
| Home self-medication | 944 (40.7%) | 561 (35.2%) | 383 (52.8%) |
| Facility Users | 1208 (52.1%) | 934 (58.6%) | 274 (37.8%) |
| **Among facility users: Choices** | **1208** | **934** | **274** |
| Pharmacy | 127 (10.5%) | 88 (9.4%) | 39 (14.2%) |
| Private clinics | 191 (15.8%) | 146 (15.6%) | 45 (16.4%) |
| Village clinics | 590 (48.8%) | 464 (49.7%) | 126 (46.0%) |
| Township health centers | 146 (12.1%) | 118 (12.6%) | 28 (10.2%) |
| County hospitals | 68 (5.6%) | 46 (4.9%) | 22 (8.0%) |
| City hospitals | 18 (1.5%) | 15 (1.6%) | 3 (1.1%) |
| Others | 68 (5.6%) | 57 (6.1%) | 11 (4.0%) |
| **Among facility users: Antibiotics Prescribed** | **1208** | **934** | **274** |
| Yes | 490 (40.6%) | 382 (40.9%) | 108 (39.4%) |
| No | 305 (25.2%) | 239 (25.6%) | 66 (24.1%) |
| Unsure | 413 (34.2%) | 313 (33.5%) | 100 (36.5%) |

*Note:* Data are presented in n (%). This table summarizes statistics on healthcare-seeking behavior and antibiotic use for treating cold or diarrhea symptoms among the surveyed families over the previous year.

**Table S6.** Household knowledge of antibiotic use (N=626).

|  | **n (%)** |
|---|---|
| **A. Household Antibiotic Knowledge Test** |  |
| As long as antibiotics can treat similar diseases, we can use friends' or families' antibiotics (T/F) (False=Correct answer) | 179 (28.6%) |
| If you recovered with antibiotics before, you can buy or ask for the same antibiotics for similar diseases (T/F) (False=Correct answer) | 129 (20.6%) |
| When should you stop using antibiotics? |  |
| When finishing all the antibiotics prescribed (correct answer) (0/1) | 31 (5.0%) |
| When feeling better (0/1) | 283 (45.2%) |
| Don't know (0/1) | 312 (49.8%) |
| **B. Household Antibiotic Recognition** |  |
| Penicillin is an antibiotic (Correct, 0/1) | 291 (46.5%) |
| Terramycin is an antibiotic (Correct, 0/1) | 193 (30.8%) |
| Yunnanbaiyao is an antibiotic (Incorrect, 0/1) | 148 (23.6%) |
| Norfloxacin is an antibiotic (Correct, 0/1) | 152 (24.3%) |
| Cough syrup is an antibiotic (Incorrect, 0/1) | 213 (34.0%) |
| Isatis root is an antibiotic (Incorrect, 0/1) | 150 (24.0%) |
| Roxithromycin is an antibiotic (Correct, 0/1) | 158 (25.2%) |
| Xiaochaihu granules is an antibiotic (Incorrect, 0/1) | 206 (32.9%) |
| Diosmectite is an antibiotic (Incorrect, 0/1) | 289 (46.2%) |
| Cephalosporin is an antibiotic (Correct, 0/1) | 236 (37.7%) |
| Insulin is an antibiotic (Incorrect, 0/1) | 272 (43.5%) |
| Metronidazole is an antibiotic (Correct, 0/1) | 144 (23.0%) |
| Niuhuangjiedu is an antibiotic (Incorrect, 0/1) | 171 (27.3%) |
| Amoxicillin is an antibiotic (Correct, 0/1) | 291 (46.5%) |

*Note:* Data are presented in n (%).

**Table S7.** OLS estimates of village doctor clinical practice quality and antibiotics use among facility users.

|  | Antibiotics Use among Facility Users (0/1) |  |  |  |  |  |  |  |
|---|---|---|---|---|---|---|---|---|
|  | (1) | (2) | (3) | (4) | (5) | (6) | (7) | (8) |
| Quality Index of Village Doctor | -0.062** | -0.066*** | -0.061*** | -0.061*** | -0.060*** | -0.061*** | -0.062*** | -0.062*** |
|  | (0.026) | (0.022) | (0.021) | (0.022) | (0.022) | (0.021) | (0.021) | (0.021) |
| Township-level Cluster SE | Y | Y | Y | Y | Y | Y | Y | Y |
| Village-level resource controls |  | Y | Y | Y | Y | Y | Y | Y |
| Provider community embeddedness |  |  | Y | Y | Y | Y | Y | Y |
| Household-level controls |  |  |  | Y | Y | Y | Y | Y |
| Individual-level controls |  |  |  |  | Y | Y | Y | Y |
| Episode-level controls |  |  |  |  |  | Y | Y | Y |
| County FE |  |  |  |  |  |  | Y | Y |
| SP FE |  |  |  |  |  |  |  | Y |
| Observation | 795 | 795 | 795 | 795 | 795 | 795 | 795 | 795 |
| R-square | 0.01 | 0.03 | 0.04 | 0.04 | 0.04 | 0.04 | 0.08 | 0.08 |

*Note:* \* p<0.1 \*\* p<0.05 \*\*\* p<0.01

**Table S8.**
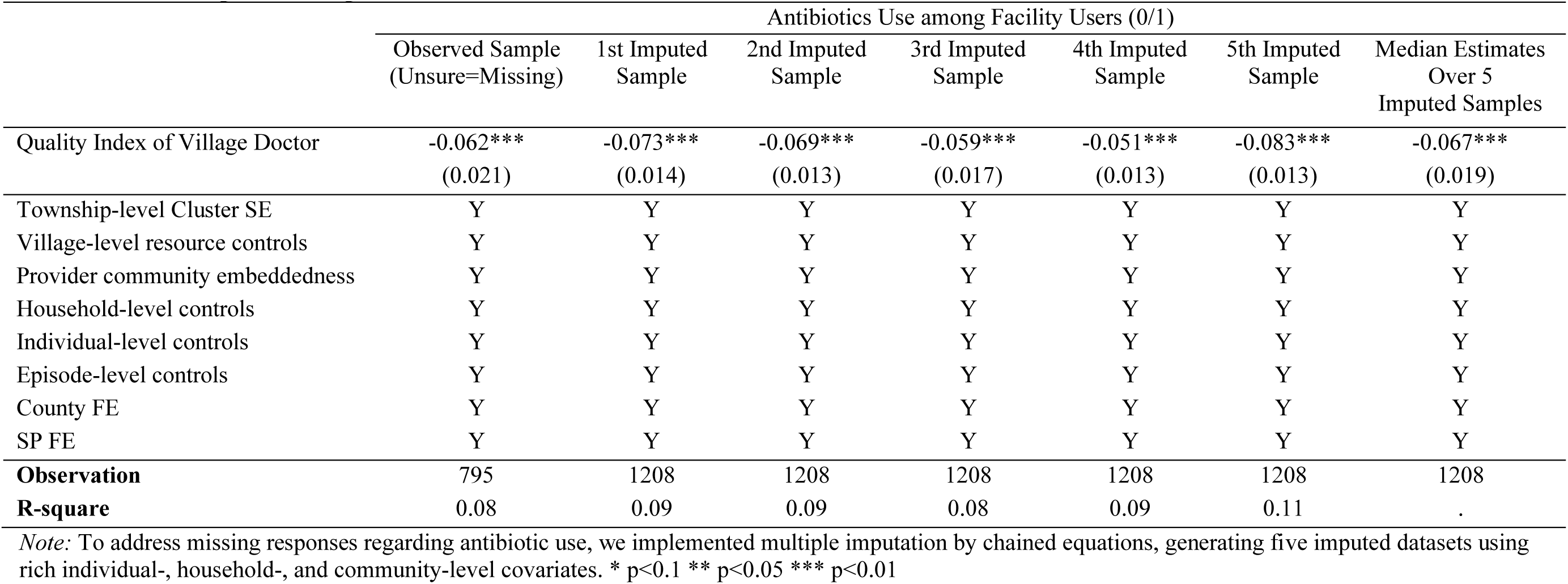
OLS estimates of village doctor clinical practice quality and antibiotic use among facility users: comparison of observed and imputed samples.

|  | Antibiotics Use among Facility Users (0/1) |  |  |  |  |  |  |
|---|---|---|---|---|---|---|---|
|  | Observed Sample (Unsure=Missing) | 1st Imputed Sample | 2nd Imputed Sample | 3rd Imputed Sample | 4th Imputed Sample | 5th Imputed Sample | Median Estimates Over 5 Imputed Samples |
| Quality Index of Village Doctor | -0.062*** | -0.073*** | -0.069*** | -0.059*** | -0.051*** | -0.083*** | -0.067*** |
|  | (0.021) | (0.014) | (0.013) | (0.017) | (0.013) | (0.013) | (0.019) |
| Township-level Cluster SE | Y | Y | Y | Y | Y | Y | Y |
| Village-level resource controls | Y | Y | Y | Y | Y | Y | Y |
| Provider community embeddedness | Y | Y | Y | Y | Y | Y | Y |
| Household-level controls | Y | Y | Y | Y | Y | Y | Y |
| Individual-level controls | Y | Y | Y | Y | Y | Y | Y |
| Episode-level controls | Y | Y | Y | Y | Y | Y | Y |
| County FE | Y | Y | Y | Y | Y | Y | Y |
| SP FE | Y | Y | Y | Y | Y | Y | Y |
| **Observation** | 795 | 1208 | 1208 | 1208 | 1208 | 1208 | 1208 |
| **R-square** | 0.08 | 0.09 | 0.09 | 0.08 | 0.09 | 0.11 | . |

*Note:* To address missing responses regarding antibiotic use, we implemented multiple imputation by chained equations, generating five imputed datasets using rich individual-, household-, and community-level covariates. \* p<0.1 \*\* p<0.05 \*\*\* p<0.01

**Table S9.** OLS estimates of village doctor clinical practice quality and antibiotic use among facility users: comparison of all facility users vs. village clinic users only.

|  | Antibiotics Use<br>among Facility Users (0/1) | Antibiotics Use<br>among Village Clinic Users only (0/1) |
|---|---|---|
| Quality Index of Village Doctor | -0.062*** | -0.055* |
|  | (0.021) | (0.032) |
| Township-level Cluster SE | Y | Y |
| Village-level resource controls | Y | Y |
| Provider community embeddedness | Y | Y |
| Household-level controls | Y | Y |
| Individual-level controls | Y | Y |
| Episode-level controls | Y | Y |
| County FE | Y | Y |
| SP FE | Y | Y |
| Observation | 795 | 406 |
| R-square | 0.08 | 0.07 |

*Note:* \* p<0.1 \*\* p<0.05 \*\*\* p<0.01

**Table S10.**
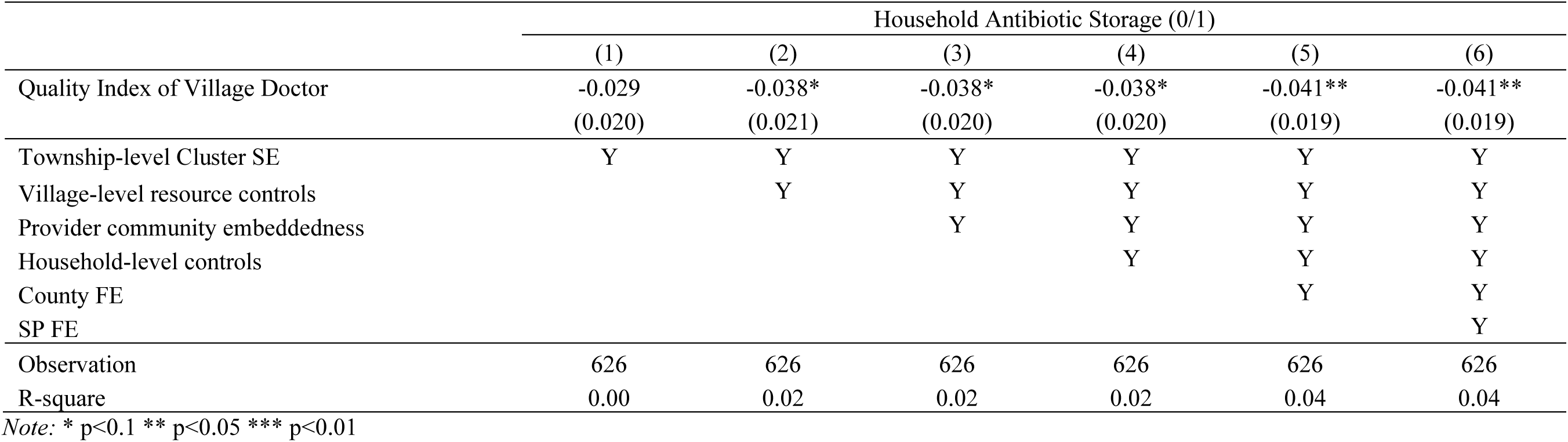
OLS estimates of village doctor clinical practice quality and household antibiotic storage.

**Table S11.** Sensitivity of IV Estimates to Violations of the Exclusion Restriction.

| **Maximum permitted violation<br>(% of reduced-form coefficient)** | **Antibiotic Receipt among Facility Users** |  | **Household Antibiotic Storage** |  |
|---|---|---|---|---|
|  | **Permitted Direct Effect<br>(δ)** | **95% UCI** | **Permitted Direct Effect<br>(δ)** | **95% UCI** |
| 0% | 0.0000 | [-0.1739, -0.0172] | 0.0000 | [-0.1741, -0.0432] |
| 10% | [-0.0080, 0.0080] | [-0.1843, -0.0084] | [-0.0091, 0.0091] | [-0.1876, -0.0348] |
| 20% | [-0.0160, 0.0160] | [-0.1948, 0.0005] | [-0.0182, 0.0182] | [-0.2012, -0.0262] |
| 30% | [-0.0240, 0.0240] | [-0.2053, 0.0095] | [-0.0273, 0.0273] | [-0.2150, -0.0174] |
| 40% | [-0.0320, 0.0320] | [-0.2159, 0.0185] | [-0.0364, 0.0364] | [-0.2289, -0.0083] |
| 50% | [-0.0400, 0.0400] | [-0.2266, 0.0276] | [-0.0455, 0.0455] | [-0.2429, 0.0010] |
| 100% | [-0.0799, 0.0799] | [-0.2809, 0.0746] | [-0.0909, 0.0909] | [-0.3142, 0.0522] |

*Note.* The table reports 95% union confidence intervals (UCI) obtained using the plausibly exogenous instrumental variables approach of Conley et al. (2012). The analysis allows the DML-constructed instrument to have a direct effect on each outcome, with the maximum permitted magnitude calibrated as a percentage of the absolute reduced-form coefficient. The estimated reduced-form coefficients are −0.0799 for antibiotic receipt among facility users and −0.0909 for household antibiotic storage. The permitted direct effects (δ) are bounded symmetrically around zero. The analyses use cross-fitted residualized variables derived from the DML-IV estimation, with standard errors clustered at the township level. Sensitivity analyses allowing for violations of the exclusion restriction showed that the 95% UCI for the negative IV estimates first included zero when the maximum permitted direct effects of the constructed instrument reached 20% of the reduced-form magnitude for antibiotic receipt and 50% for household antibiotic storage.

**Table S12.**
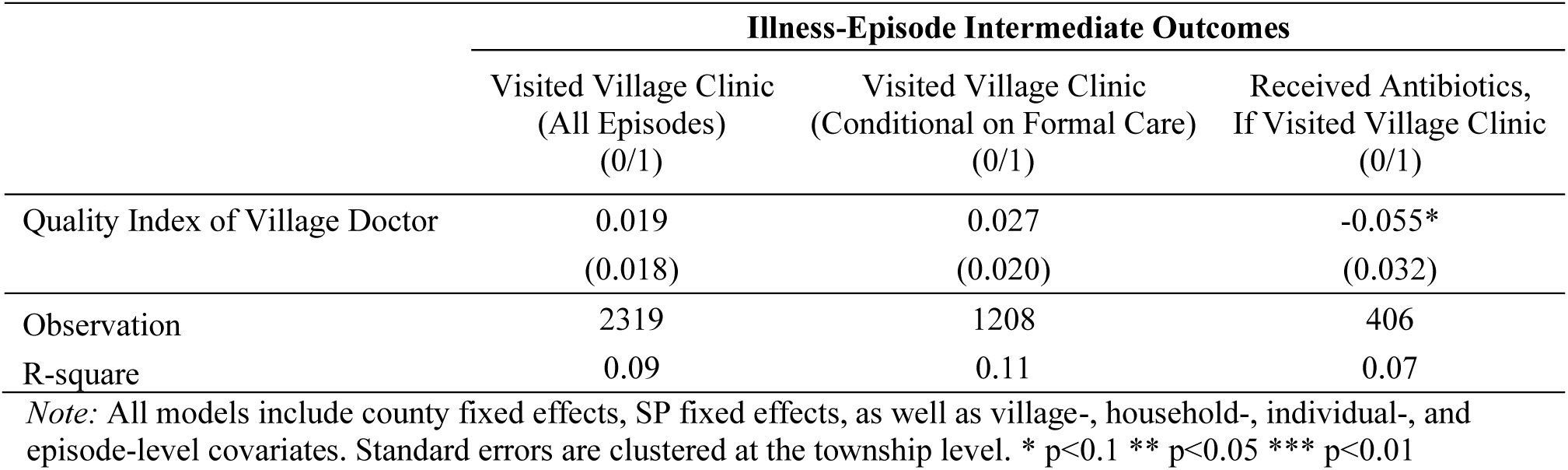
Village doctor clinical practice quality and intermediate outcomes.

**Table S13.** Village doctor clinical practice quality and intermediate outcomes.

|  | **Household-level Intermediate Outcomes** |  |
|---|---|---|
|  | Household Stated Preferences for Village Clinic Care (0/1) | Household Perceived Quality of Village Clinics |
| **Panel A. Objective Quality** |  |  |
| Objective Quality of Village Doctor | 0.006 | 0.036 |
|  | (0.018) | (0.043) |
| Observation | 624 | 624 |
| R-square | 0.07 | 0.07 |
| **Panel B. Perceived Quality** |  |  |
| Perceived Quality of Village Doctor | 0.081*** |  |
|  | (0.015) |  |
| Observation | 622 |  |
| R-square | 0.10 |  |

*Note:* All models include county fixed effects, SP fixed effects, as well as village-, household-, individual-, and episode-level covariates. Standard errors are clustered at the township level. \* p<0.1 \*\* p<0.05 \*\*\* p<0.01

**Table S14.** Village doctor clinical practice quality and intermediate outcomes.

|  | **Household-level Intermediate Outcomes** |  |
|---|---|---|
|  | Household Antibiotic Use Knowledge Index | Household Antibiotic Recognition Index |
| Quality Index of Village Doctor | -0.025 | -0.003 |
|  | (0.041) | (0.038) |
| Observation | 626 | 626 |
| R-square | 0.08 | 0.08 |

*Note:* All models include county fixed effects, SP fixed effects, as well as village-, household-, individual-, and episode-level covariates. Standard errors are clustered at the township level. \* p<0.1 \*\* p<0.05 \*\*\* p<0.01

### C. Description of DML-IV Framework

The double/debiased machine learning (DML) framework supports flexible estimation of causal effects in settings where the functional form of confounding is unknown and potentially nonlinear by incorporating a broad set of machine learning methods (Ahrens et al., 2024, 2026; Chernozhukov et al., 2017, 2018). Conventional estimators such as OLS often impose linear functional forms, which may not be sufficiently flexible to capture complex confounding, whereas fully nonparametric estimators quickly become infeasible in high-dimensional settings with large and complex datasets (i.e., the “curse of dimensionality”).

Machine learning methods have become increasingly popular for addressing these challenges (Athey & Imbens, 2019; Mullainathan & Spiess, 2017). But many existing applications rely primarily on lasso-based regularization procedures (Belloni et al., 2012; Chernozhukov et al., 2015), which may be ill-suited when approximate sparsity is weak, as in our setting. In contrast, DML supports a broad class of supervised learning algorithms, such as random forests and gradient boosting. By combining these flexible learners with causal estimators from traditional econometric models, DML enables more flexible estimation of causal effects while accommodating complex, high-dimensional data.

The DML estimators operate in two steps. The first step uses cross-fitting to estimate conditional expectation functions (CEFs) with flexible, potentially nonparametric learners. In the second step, both the outcome and the endogenous regressor are “residualized” by subtracting their respective CEF estimates from the first step. These residuals are then used for a second-stage estimation based on Neyman-orthogonal scores. Further technical details are provided in Chernozhukov et al. (2018) (Chernozhukov et al., 2018). Standard errors for DML estimators are obtained in the usual way from the second-stage regression using the residualized variables. Thus, the standard error corrections available for linear regression, including clustered standard errors (used here at the township level), are directly applicable (Ahrens et al., 2024).

The first-stage cross-fitting involves randomly splitting the sample into *K* approximately equal folds. For each fold, the CEFs are estimated using only observations outside that fold, and the models are then used to generate out-of-sample predictions for observations within the fold. This procedure is repeated for all *K* folds so that every observation receives predictions from models trained on disjoint data, and the resulting CEF estimates are then averaged across folds. This procedure mitigates overfitting and ensures that first-stage estimation errors are approximately independent of the residuals used in the second stage (Chernozhukov et al., 2018). Because the first-stage CEFs can be estimated with a wide range of supervised learning algorithms, DML provides a flexible framework for estimating causal effects in high-dimensional, nonlinear settings where traditional parametric methods may fall short.

#### DML Flexible Partially Linear IV Model

In this study, we implemented the DML flexible partially linear IV model, which allowed us to approximate an optimal instrument from a large set of potential instruments using a broad class of machine learning techniques. We used the 18 provider characteristics as the baseline instrument set and allowed for flexible function approximation by including first-order interaction terms and second-order polynomial terms. Control variables followed a similar setup, with baseline variables, interactions, and square terms included in CEF estimation.

For the first-stage cross-fitting of the CEFs, we set *K*=5 for cross-fitting and implemented cluster-dependent cross-fitting by assigning folds at the township level to respect within-cluster dependence (Ahrens et al., 2024). Standard errors were clustered at the same level.

In terms of the selection of machine learners, we incorporated multiple supervised learning algorithms with varying parameters, including OLS, cross-validated ridge, random forest, gradient boosting, and neural network. Since it is often difficult to know ex ante which algorithm would perform best in a given application, we applied a newly proposed “stacking” approach to combine different machine learners into a single meta-learner (Ahrens et al., 2023, 2025). This stacking ensemble approach assigns weights (between 0 and 1) to each learner based on out-of-sample performance, effectively down-weighting poorly performing or misspecified learners and placing greater weight on those with lower prediction error. Research has shown that such stacking procedures are robust to ill-chosen learners and typically outperform individual algorithms (Ahrens et al., 2023, 2025).

In this setting, gradient boosting received the largest stacking weights (approximately 0.95-0.99), indicating its dominant role in approximating the optimal instrument. This likely reflects its ability to flexibly capture nonlinearities and interactions among many individually weak but jointly informative predictors of provider performance, making it particularly well suited for approximating an optimal instrument for real-world clinical practice quality.

